# The neonatal gut microbiota: a role in the encephalopathy of prematurity

**DOI:** 10.1101/2023.09.12.23295409

**Authors:** Kadi Vaher, Manuel Blesa Cabez, Paula Lusarreta Parga, Justyna Binkowska, Gina J van Beveren, Mari-Lee Odendaal, Gemma Sullivan, David Q Stoye, Amy Corrigan, Alan J Quigley, Michael J Thrippleton, Mark E Bastin, Debby Bogaert, James P Boardman

## Abstract

Preterm birth associates with atypical brain development and neurocognitive impairment. The gut microbiome is implicated in neurobehavioural outcomes of typically developing infants and children, but its relationship with neurodevelopment in preterm infants is unknown. We characterised the faecal microbiome in a cohort of 147 neonates enriched for very preterm birth (<32 weeks’ gestation) using 16S-based and shotgun metagenomic sequencing. Delivery mode had the strongest association with preterm microbiome shortly after birth; low birth gestational age, infant sex and antibiotics significantly associated with microbiome composition at NICU discharge. Thereafter, we integrated these data with term-equivalent structural and diffusion brain MRI. Bacterial community composition associated with MRI features of encephalopathy of prematurity. Particularly, abundances of *Escherichia coli* and *Klebsiella* spp. correlated with microstructural parameters in deep and cortical grey matter. Metagenome functional capacity analyses suggested that these bacteria may interact with brain microstructural development via tryptophan and propionate metabolism. This study indicates that gut microbiome associates with brain development following preterm birth.

## INTRODUCTION

Globally, preterm birth, defined as birth before 37 weeks of gestation, affects around 10% of pregnancies^1^. People born preterm are at an increased risk for atypical brain development, termed encephalopathy of prematurity (EoP)^2^, which can lead to cerebral palsy, neurodevelopmental and cognitive impairments, autism, and psychiatric disorders^3^. There are no treatments for EoP, partly because the mechanisms linking preterm birth with altered cerebral development are incompletely understood.

The second and third trimesters of pregnancy are crucial periods in brain development. During this time, preterm birth and its co-exposures and -morbidities impose a risk of injury and dysmaturation to the developing brain, leading to disturbances in regional brain growth, diffuse white matter disease, abnormal cortical and deep grey matter (dGM) development, and structural dysconnectivity^4^. These features of EoP are apparent on structural and diffusion magnetic resonance imaging (MRI) in the neonatal period^4^ and, because they are associated with subsequent neurocognitive development^5–8^, they serve as intermediate phenotypes to investigate the upstream determinants of brain development.

Fundamental neurodevelopmental processes occurring in early life coincide with the acquisition and progression of the gut microbiota. Evidence from preclinical and human observational studies implicates the gut microbiome in modulating neural functions via the microbiota-gut-brain axis^9,10^. Specifically, the rapid parallel development of the brain and the gut microbiota in early life has led to the hypothesis of ‘nested sensitive periods’ whereby brain development interacts with gut microbiota development to shape cognition and behaviour^11,12^. The hypothesis has gained traction from a growing body of literature reporting associations between gut microbiota features and cognitive, language, motor, and socio-emotional development in childhood^13^.

Preterm infants may be particularly vulnerable to disruptions in the microbiota-gut-brain axis due to altered microbiota development, which can arise from the early exposure of the immature gastrointestinal tract to microbial colonisation^14,15^. Although the general pattern of microbiota development in the first months of life appears similar in term and preterm infants^16–18^, the preterm infant gut has lower bacterial diversity and abundances of essential microbes like *Bifidobacterium*, and higher levels of opportunistic pathogens such as *Klebsiella, Enterobacter, Enterococcus* and *Staphylococcus*^14^. This may be a result of routine exposure to potent modifiers of the pioneering microbiota, including maternal and neonatal antibiotic treatments^19^, and variable nutritional exposures^20,21^ during the first months of life in a neonatal intensive care unit (NICU) setting. However, there are discrepancies between studies about the effect size and direction of these modifiers in preterm neonates^22^, which leaves considerable uncertainty about the importance of specific clinical variables for shaping microbiota development following preterm birth.

Although the preterm population has a high burden of neurocognitive impairment and alterations in the gut microbiota, only a few recent studies have investigated gut microbiota in direct relation to preterm infant neurodevelopment^23–27^ or overt parenchymal brain injuries^28^. Though most of these studies have been small and directions of effects vary, there is some consensus that abundances of *Bifidobacteriaceae*, *Enterococcaceae, Enterobacteriaceae* (*Escherichia/Shigella, Enterobacter, Klebsiella*), *Clostridium,* and *Veillonella* may correlate with outcomes. However, to our knowledge, no studies have investigated the preterm microbiota in association with EoP. This question is important because EoP is the prevailing form of brain dysmaturation after preterm birth and the gut microbiota is intrinsically modifiable by mode of feeding and enteral supplements, which offer potential new avenues for perinatal neuroprotection.

We investigated the microbiota-gut-brain axis by integrating data from microbiota profiling and multimodal brain MRI. We aimed to characterise neonatal gut microbiota profiles in a richly-phenotyped cohort of term and preterm neonates at birth and at NICU discharge; to determine the most influential clinical drivers of the preterm microbiota during NICU care; and to link gut microbiota diversity and community composition with MRI features of EoP.

## RESULTS

### SAMPLE CHARACTERISTICS

The gut microbiome was sampled at two timepoints (TP1: meconium, and TP2: a faecal sample prior to discharge from NICU) in very preterm infants born < 32 completed weeks of gestation, and at TP1 in term-born controls, who were recruited to the Theirworld Edinburgh Birth Cohort (TEBC)^29^. Clinical and demographic characteristics of the study group are shown in Table 1 and Table S1; see Figure S1 for flowchart.

**Table 1.**
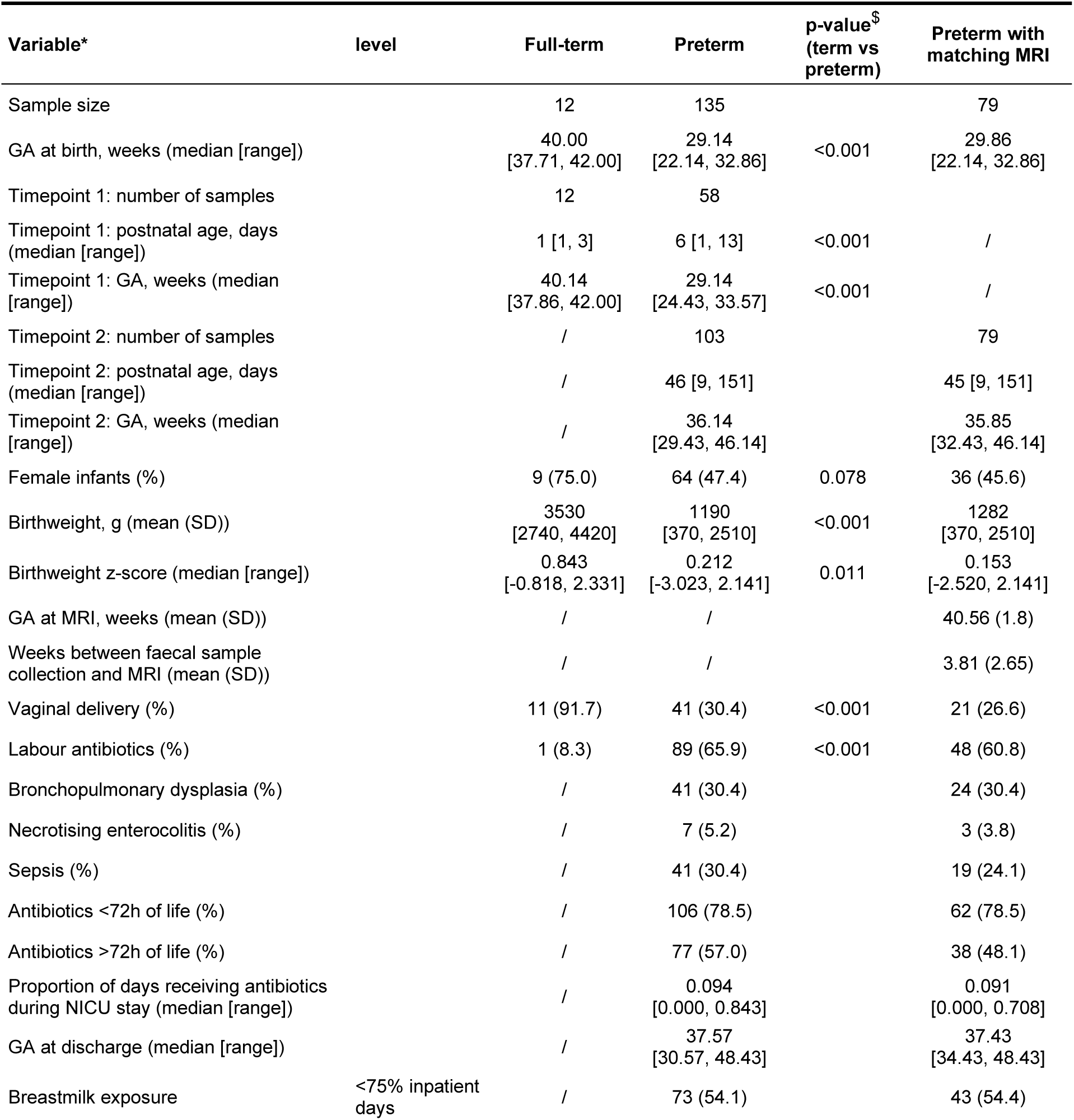
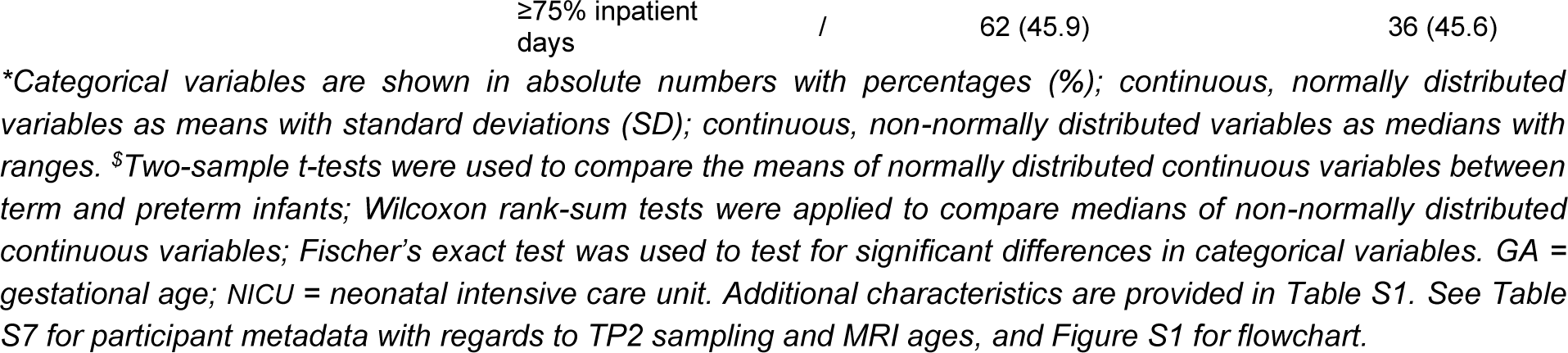
Baseline characteristics of the study group.

### OVERVIEW OF MICROBIOTA PROFILES

We first characterised neonatal intestinal microbiome profiles using 16S ribosomal RNA (rRNA) gene sequencing in 12 term and 58 preterm infants at TP1 and in 103 preterm infants at TP2. Shotgun metagenomic sequencing data was available for 23 preterm infants at TP1 and 97 preterm infants at TP2. Throughout the paper, 16S-based data is described at amplicon sequence variant (ASV)-level, while shotgun taxonomic data is described at species-level.

The majority of TP1 samples were dominated by an ASV from the genus *Staphylococcus*, but some had high relative abundances of ASVs belonging to genera *Streptococcus*, *Escherichia/Shigella, Enterococcus,* or *Klebsiella* (Figure 1A). The most abundant species (shotgun data) in the subset of TP1 samples were *Escherichia coli, Enterococcus faecalis, Staphylococcus epidermidis* and *S. haemolyticus,* and *Raoultella planticola* (Figure S2A). At TP2, most samples had high relative abundances of ASVs belonging to *Bifidobacterium, Enterobacteriaceae* or *Escherichia/Shigella,* whilst some had high relative abundances of a *Klebsiella* ASV (Figure 1A). Shotgun sequencing showed similar profiles, but allowed better species-level resolution, with high abundances of *Bifidobacterium* spp. (*B. breve, B. longum, B. dentium*), *E. coli, E. faecalis*, or *Klebsiella* spp. (*K. pneumoniae, K. oxytoca*; Figure S2A).

**Figure 1.**
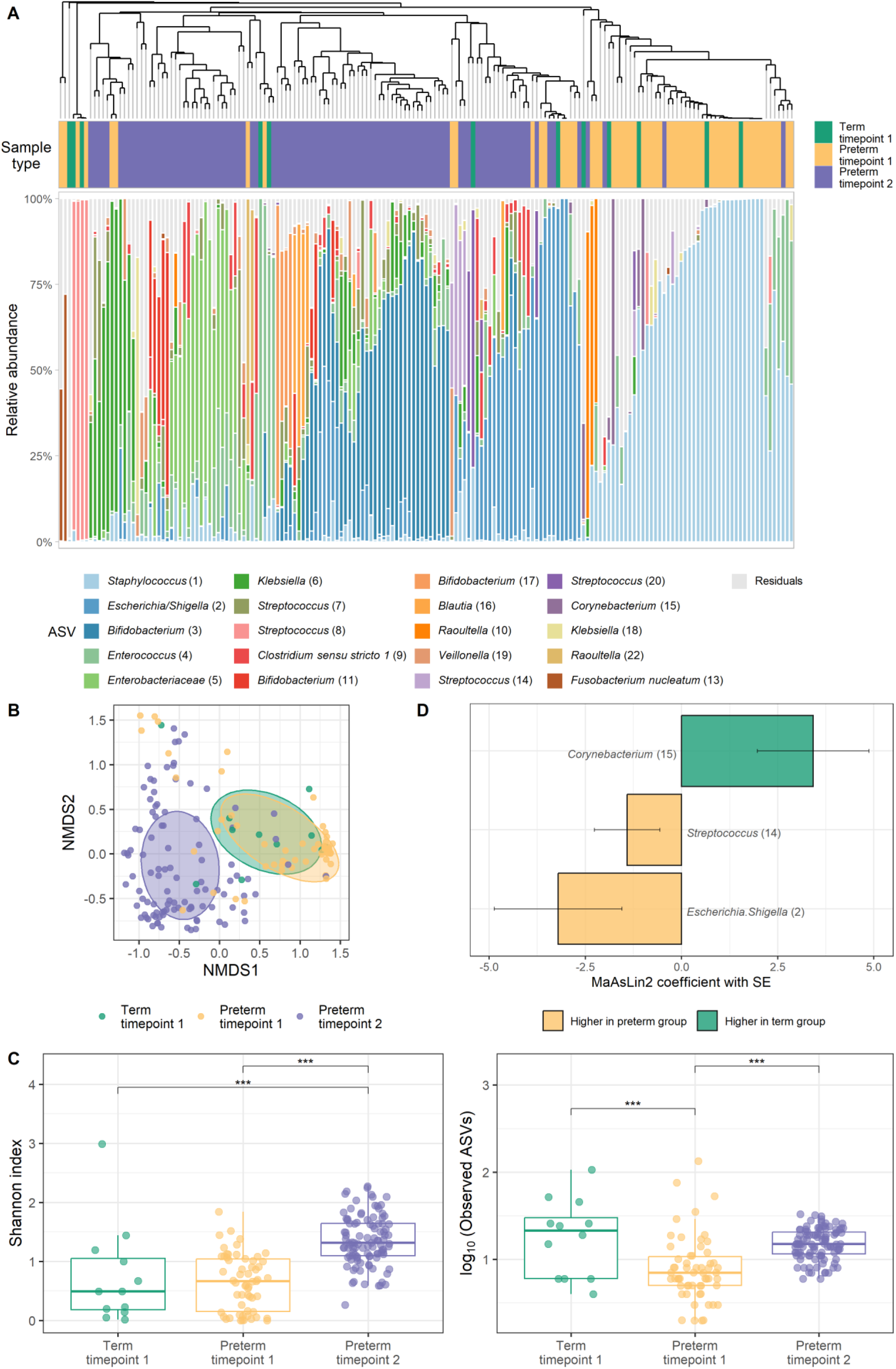
Overview of microbiota profiles in neonates based on 16S rRNA gene sequencing. (A) Relative abundances of the 20 most abundant amplicon sequence variants (ASVs) identified across the dataset are visualised per sample, with all other ASVs grouped together as residuals. Samples are ordered based on hierarchical clustering of the Bray-Curtis dissimilarity matrix using average linkage (see dendrogram). (B) Non-metric multidimensional scaling (NMDS) plot based on Bray-Curtis dissimilarity between samples; data points and ellipses are coloured by sample type. The ellipses denote the standard deviation of data points belonging to each sample type, with the centre points of the ellipses calculated using the mean of the coordinates per group. (C) Microbiota alpha diversity measured by Shannon index (left) and observed ASVs (right). *** indicates q < 0.001 in pairwise comparisons using *emmeans* following linear mixed effects model comparing alpha diversity indices between the groups and timepoints. (D) Differentially abundant ASVs in association with preterm status at timepoint 1. Bar plots depict MaAsLin2-analysis results. ASVs present with at least 1% of abundance in at least 5% of samples were analysed (10 ASVs) and significant results are shown (BH corrected *p* < 0.25 as default). Lengths of the bars correspond with the MaAsLin2-model coefficient, which relates to the strength of the association. Error bars indicate the standard error (SE) of the model coefficient. MaAsLin2 models were adjusted for postnatal age at sampling. Sample sizes: term timepoint 1 = 12, preterm timepoint 1 = 58, preterm timepoint 2 = 103. See Figure S2 for overview of microbiome profiles in preterm neonates arising from shotgun sequencing.

Collectively, we observed a marked shift in the preterm gut microbiota community composition between birth and term-equivalent age (TEA) when analysed at ASV- or species-level (permutational analysis of variance [PERMANOVA] R^2^ = 14.59%, p = 9.99 × 10^−4^ [Figure 1B]; R^2^ = 3.31%, p = 0.002 [Figure S2C], respectively). Differences in the microbiota between the two timepoints in preterm infants were also reflected in increasing alpha diversity (Figure 1C; linear mixed effects model; F_2,112.86_ = 47.244, q = 2.48 × 10^−15^ for Shannon index [pairwise comparison: q = 4.00 × 10^−14^]; F_2,151.99_ = 22.876, q = 2.06 × 10^−9^ for observed ASVs [pairwise comparison: q = 6.88 × 10^−9^]). Microbiome functional capacity captured by gut-metabolic modules (GMM; Figure S2B), similarly to taxonomic composition, also differed between the two timepoints in preterm infants (PERMANOVA R^2^ = 4.17%, p = 9.99 × 10^−4^; Figure S2D).

Comparison of preterm infants with a small sample of term-born controls at TP1 revealed weak evidence for a small difference in bacterial community composition at ASV-level (R^2^ = 2.75%, p = 0.082; Figure 1B). This primarily manifested in lower bacterial richness (observed ASVs, q = 7.75 × 10^−5^; Figure 1C) and higher relative abundances of ASVs belonging to *Escherichia/Shigella* (q = 0.189) and *Streptococcus* (q = 0.223) genera in the preterm group, while term infants had higher abundance of a *Corynebacterium* ASV (q = 0.131; Figure 1D).

### COVARIATES SHAPING PRETERM INFANT GUT MICROBIOME

We then sought to identify perinatal covariates associated with microbiota communities in our preterm cohort, focussing on variables known from literature to associate with microbiota composition in infancy (gestational age [GA] at birth^17,18^, age at sampling^18,30,31^, birthweight^20^, delivery mode^32–34^, antibiotics^35–37^, breastmilk exposure^20,30,38^), and common preterm neonatal co-morbidities (sepsis^37,39^, necrotising enterocolitis [NEC]^40^, bronchopulmonary dysplasia [BPD]^41,42^). Although sex is not often investigated in association with early life gut microbiota development and previous studies report mixed findings to the extent that sex associates with infant microbiota^30,43,44^, we included this variable because boys and girls differ in susceptibility to mortality and major morbidities following preterm birth^45^.

In univariable models, ASV-level bacterial community composition at TP1 significantly associated with mode of delivery, birthweight z-score, and postnatal age at sample collection (Figure 2A left panel). Using shotgun sequencing, the different perinatal factors had relatively stronger correlations with species-level community composition, though most did not remain significant after adjustment for multiple comparisons (Figure 2A middle panel).

**Figure 2.**
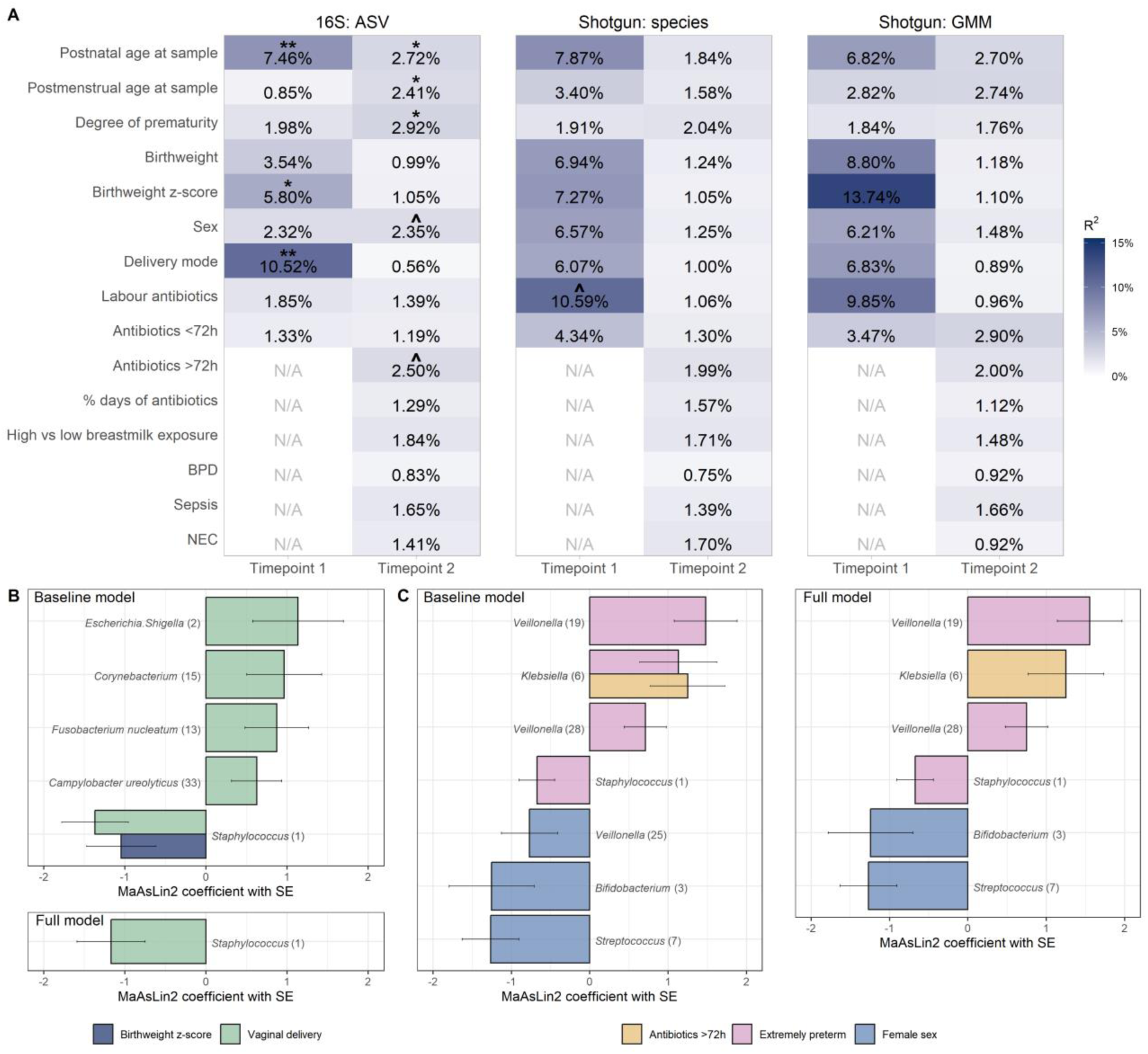
Covariates associated with preterm infant gut microbiota. (A) Univariable PERMANOVA results showing the association between perinatal variables and the gut bacterial community composition at each timepoint and for each data type: left panel = ASV from 16S rRNA sequencing, middle panel = species from shotgun sequencing, right panel = Gut metabolic modules (GMM) calculated from KEGG orthologs arising from shotgun sequencing. The variance explained is estimated for each variable independently and is indicated by a percentage/blue shades. Significance of PERMANOVA was based on 1000 permutations and was adjusted for multiple comparisons using the Benjamini-Hochberg (BH) method; asterisks denote statistical significance (^*q* ≤ 0.1, \**q* ≤ 0.05, \*\**q*≤ 0.01). (B, C) Differentially abundant ASVs in association with perinatal factors at timepoint 1 (B) and 2 (C). Bar plots depict MaAsLin2-analysis results. ASVs present with at least 1% of abundance in at least 5% of samples were analysed (14 ASVs for timepoint 1 and 21 for timepoint 2) and significant results are shown (BH corrected *p* < 0.25 as default). Bars are coloured according to the covariate they are associated with. Lengths of the bars correspond with the MaAsLin2-model coefficient, which relates to the strength of the association. Error bars indicate the standard error (SE) of the model coefficient. In baseline models, we adjusted for postnatal age (timepoint 1), or GA at birth and sample collection (timepoint 2); in full adjusted models, all covariates with q-value < 0.1 from univariable PERMANOVAs were tested simultaneously. Here, GA at birth was dichotomised to group the infants into extremely (GA at birth < 28 completed weeks) and very (GA at birth < 32 completed weeks) preterm. Sample sizes for 16S rRNA sequencing: preterm timepoint 1 = 58, preterm timepoint 2 = 103; sample sizes for shotgun sequencing: preterm timepoint 1 = 23, preterm timepoint 2 = 97. See Tables S2-S5 for detailed MaAsLin2 results, and Table S6 for alpha diversity associations.

At TP2, we observed the strongest associations for ASV-level bacterial community composition with the degree of prematurity, followed by postnatal age and GA at sample collection, antibiotic exposure, and sex (Figure 2A left panel). Using shotgun sequencing, none of the tested covariates were statistically significantly associated with community composition, though ranking of the effect sizes was similar (Figure 2A middle panel).

Using 16S-based ASV-level data (Tables S2-3), Microbiome Multivariate Association with Linear Models (MaAsLin2^46^) revealed that delivery mode (Figure 2B top panel) associated, among others, with the abundances of *Escherichia/Shigella* (q = 0.172) and *Staphylococcus* (q = 0.023). Birthweight z-score negatively correlated with the abundance of *Staphylococcus* (q = 0.217), but not when adjusting for delivery mode (Figure 2B bottom panel). At TP2 (Figure 2C), the faecal samples of extremely compared to very preterm infants had higher relative abundances of different *Veillonella* ASVs (q range 0.008 – 0.078) and *Klebsiella* (q = 0.160), and lower levels of *Staphylococcus* (q = 0.037; Figure 2C left panel). Male and female infants differed in the relative abundances of *Veillonella* (q = 0.216)*, Bifidobacterium* (q = 0.171), and *Streptococcus* (q = 0.011), and antibiotic-exposed infants had higher relative abundance of *Klebsiella* (q = 0.110). The main results of the full mutually adjusted model (Figure 2C right panel) paralleled those of the baseline model. Species-level analyses from shotgun sequencing showed similar top hits to those observed with ASVs from 16S (Tables S4-5), including the higher abundance of *Veillonella parvula* in extremely preterm infants and lower levels of *Streptococcus vestibularis* in females at TP2.

Bacterial alpha diversity minimally correlated with the perinatal covariates (Table S6): no significant correlations were found at TP1, whilst at TP2, bacterial richness correlated positively with age at sampling (postnatal and GA), and birthweight z-score, and bacterial richness was higher in extremely compared to very preterm infants, and in infants diagnosed with NEC.

Complementary analysis of the functional capacity of the microbiome using the GMMs (Figure 2A right panel) revealed nominally significant associations for birthweight z-score and labour antibiotics at TP1, though these relationships did not remain significant after adjustment for multiple comparisons. At TP2, none of the covariates tested had statistically significant associations with community composition at the functional GMM level.

### GUT MICROBIOTA ASSOCIATIONS WITH MRI FEATURES OF ENCEPHALOPATHY OF PREMATURITY

Following characterisation of the preterm infant microbiota, we investigated associations between the gut microbiome and MRI biomarkers of EoP at TEA in 79 infants; brain MRI scans were conducted, on average, 3.81 weeks after the collection of TP2 sample (Table S7).

We first sought to reduce the multidimensionality of the data into a meaningful set of variables capturing the variation in the microbiota compositional data. We extracted four principal coordinates (PCo) calculated from ASV-level Bray-Curtis dissimilarity matrix; these together explained 40.9% of variance in the microbiota community composition data. Correlation analysis between the relative abundance of ASVs and the four PCo-s revealed that PCo1 mainly indicated lower relative abundances of *Bifidobacterium* and *Cutibacterium*, and, though with a weaker correlation coefficient, higher abundances of *Staphylococcus* and a set of *Enterobacteriaceae*; PCo2 indicated lower relative abundances of *Escherichia/Shigella* and higher abundances of an unidentified ASV in *Enterobacteriaceae* family; PCo3 mainly indicated lower abundances of *Klebsiella* and, to lesser extent, higher abundances of *Enterobacteriaceae*; and PCo4 indicated lower abundances of *Enterococcus* and interestingly, but to a lesser extent, both higher and lower abundances of different *Bifidobacterium* ASVs (Figure 3). These PCo-bacteria correlations were confirmed using metagenomic sequencing (Figure S3), though simultaneously providing better species resolution. Notably, PCo2 indicated higher abundances of *K. oxytoca*, suggesting a species-specification to the unnamed *Enterobacteriaceae* ASV from 16S-based sequencing. From 16S data, we calculated the Shannon index and number of observed ASVs as two complementary measures of alpha diversity. The PCo-s were orthogonal and showed very weak rank correlations with one another (Spearman ρ range 0.03-0.19), suggesting that each of them captures an independent aspect of the variance in gut bacterial community composition. The two alpha diversity indices were, as expected, moderately correlated with one another (Spearman ρ = 0.63); furthermore, observed ASVs had a moderate negative correlation with PCo1 (Spearman ρ = −0.49).

**Figure 3.**
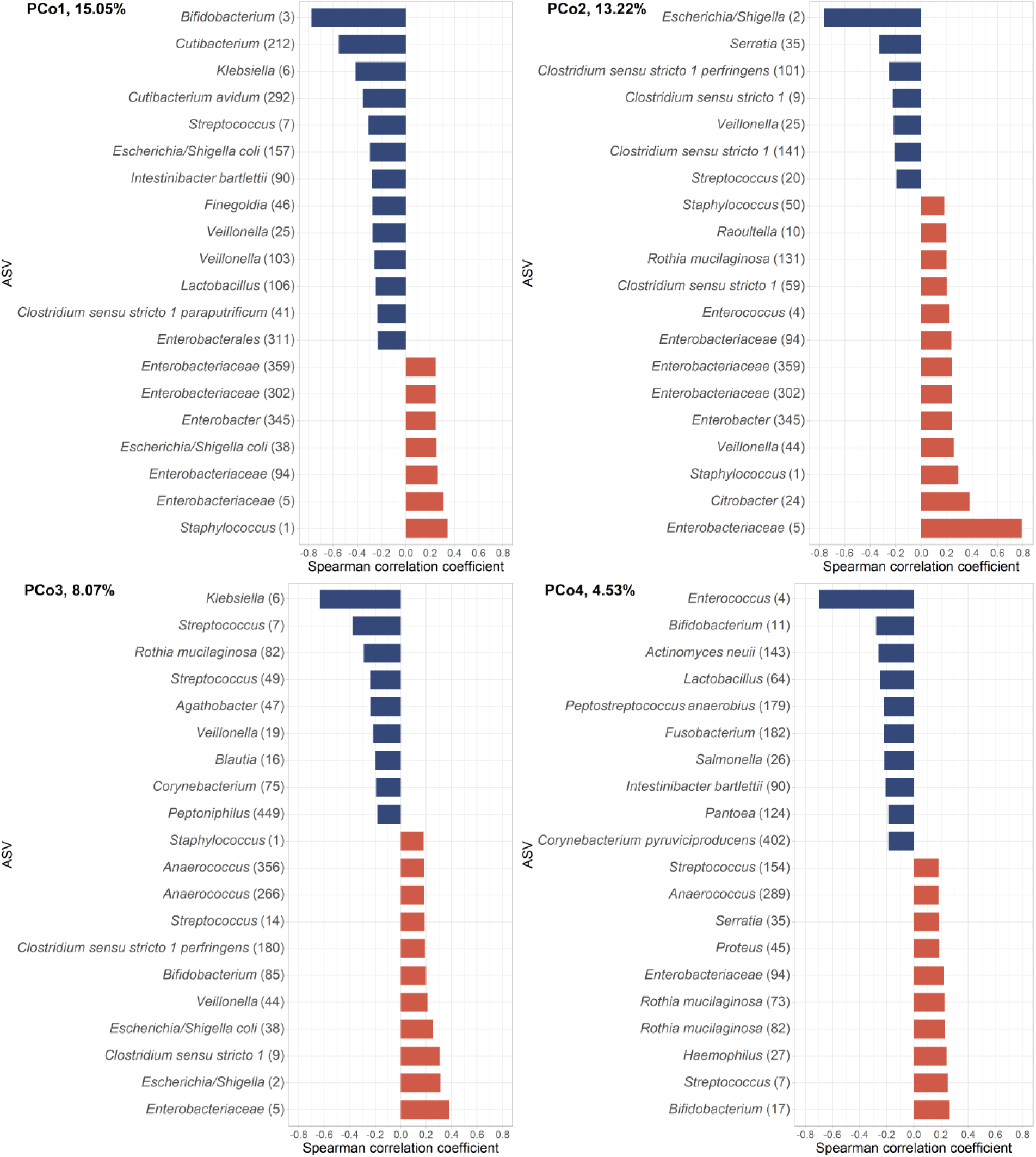
Dimensionality reduction of the microbiota community composition data. Bacterial ASV correlations with the first four orthogonal principal coordinates (PCo), showing the top 20 strongest correlations for each PCo. The % refers to the variance explained by each of the PCos. Red indicates positive and blue negative correlations between the PCo-s and ASVs. Sample size n=79 (linked MRI and microbiome data). See Figure S3 for bacterial species’ (shotgun sequencing) correlations with the PCo-s.

The four beta-diversity PCo-s and two alpha diversity indices were then used as main predictors of interest in studying the relationships between gut microbiota and MRI biomarkers of EoP. We focussed on whole-brain imaging metrics capturing brain size (tissue volumes), microstructure derived from diffusion tensor imaging (DTI; fractional anisotropy [FA], radial diffusivity [RD]) and neurite orientation dispersion and density imaging (NODDI; neurite density index [NDI], orientation dispersion index [ODI], isotropic volume fraction [ISO]), and cortical morphometry (gyrification index, thickness, sulcal depth, curvature, surface area). For contextualisation of the image features in respect to GA at birth and at scan, please see Table S8.

PCo1 negatively correlated with total brain tissue and absolute white matter volume, and positively with relative dGM volume (Figure 4A). The incremental R^2^ upon adding the PCo1 to a null model was 2.6% for total brain tissue, 5.4% for white matter, and 8.3% for dGM relative volume. There was also a nominally significant association between relative cortical volume and PCo2 (incremental R^2^ 2.6%). However, no volumetric association remained statistically significant after multiple comparison adjustment.

**Figure 4.**
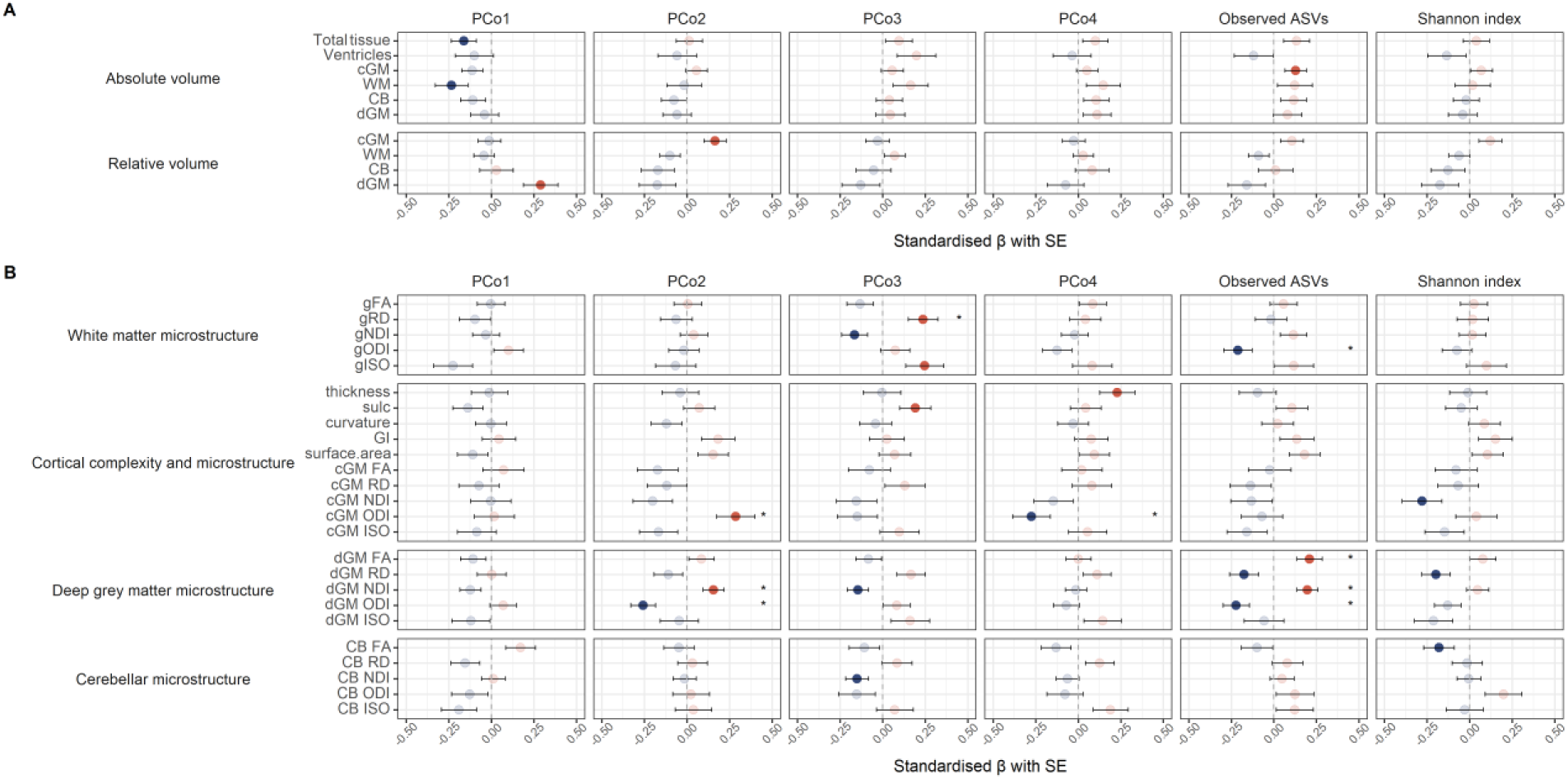
Microbiota associations with MRI features of encephalopathy of prematurity. (A) Regression results for brain volumetric measures. (B) Regression results for brain microstructural measures. Models are adjusted for gestational age at birth and at scan; microbiota PCo-s and alpha diversity metrics were adjusted for gestational age at sampling via linear regression, retaining the residuals. Full colour points indicate nominal p-value < 0.05; asterisks (*) indicate Benjamini-Hochberg (BH)-method-adjusted p-value < 0.25. Red indicates positive and blue negative associations. Relative volumes are calculated by normalising to total tissue volume (the sum of the volumes of cortical grey matter, white matter, deep grey matter, cerebellum, brainstem, hippocampi and amygdalae). FA = fractional anisotropy; RD = radial diffusivity; NDI = neurite density index; ODI = orientation dispersion index; ISO = isotropic volume fraction; cGM = cortical grey matter; dGM = deep grey matter, CB = cerebellum; sulc = sulcal depth; GI = gyrification index, g = general factor. Sample sizes (total n=79): volumetric and cortical structural complexity analysis = 76, white matter microstructure analysis = 74, and cortical and deep grey matter and cerebellar microstructural diffusion analysis = 74. See Table S8 for contextualisation of the image features in respect to GA at birth and at scan, Tables S9-S10 for ASV- and species-level MaAsLin2 results, and Figure S5 for representative brain maps.

In contrast, there were statistically significant associations between the microbiota and dMRI features of EoP after correction for multiple tests (Figure 4B). PCo2 associated with dGM microstructure (incremental R^2^ was 6.4% for ODI and 2.3% for NDI) and ODI in the cortex (incremental R^2^ 7.8%); PCo3 associated with measures of global white matter microstructure (gRD, gNDI and gISO; incremental R^2^ was 5.6%, 2.7% and 6.0%, respectively); and PCo4 associated with cortical complexity (thickness; incremental R^2^ 5.2%) and microstructure (ODI; incremental R^2^ 7.5%). Microbiota richness (number of observed ASVs) also associated with dGM microstructure (incremental R^2^ was 3.9%, 3.4%, 4.3% and 2.7%, for FA, NDI, ODI and RD, respectively) as well as ODI in the white matter (incremental R^2^ 4.3%).

The statistically significant microbiota-brain associations after FDR correction (except for the correlation between observed ASVs and gODI), remained significant in sensitivity analyses where we adjusted for birthweight z-score and sex, and excluded infants with NEC (data not shown).

PCo-s capture complex patterns of variation within bacterial communities; therefore, to better understand how specific bacterial biomarkers may be related to brain features, we performed post-hoc analysis using MaAsLin2 for those MRI features that showed statistically significant associations with gut microbiota in PCo- and alpha diversity-based analyses. These results were partially in line with those obtained using the microbial community PCo-s (Figure 5; Table S9). *Bifidobacterium*, the strongest driver of the PCo1, was not significantly associated with any of the brain microstructural measures tested. In line with the PCo2-dGM microstructure findings, *Escherichia/Shigella*, the strongest negative driver of PCo2, showed significant associations with FA (q = 0.204), NDI (q = 0.004), and ODI (q = 0.061) in dGM. Also in line, *Enterobacteriaceae*, the strongest positive driver of PCo2, correlated significantly with dGM ODI (q = 0.025). *Klebsiella*, the strongest negative driver of PCo3, was ranked at the top of the list of associations with gRD (q = 0.328), and was interestingly significantly associated with NDI (q = 0.089) and ODI (q = 0.162) in dGM. In contrast, *Enterococcus*, the strongest negative driver of PCo4, was not significantly associated with any of the MRI markers.

**Figure 5.**
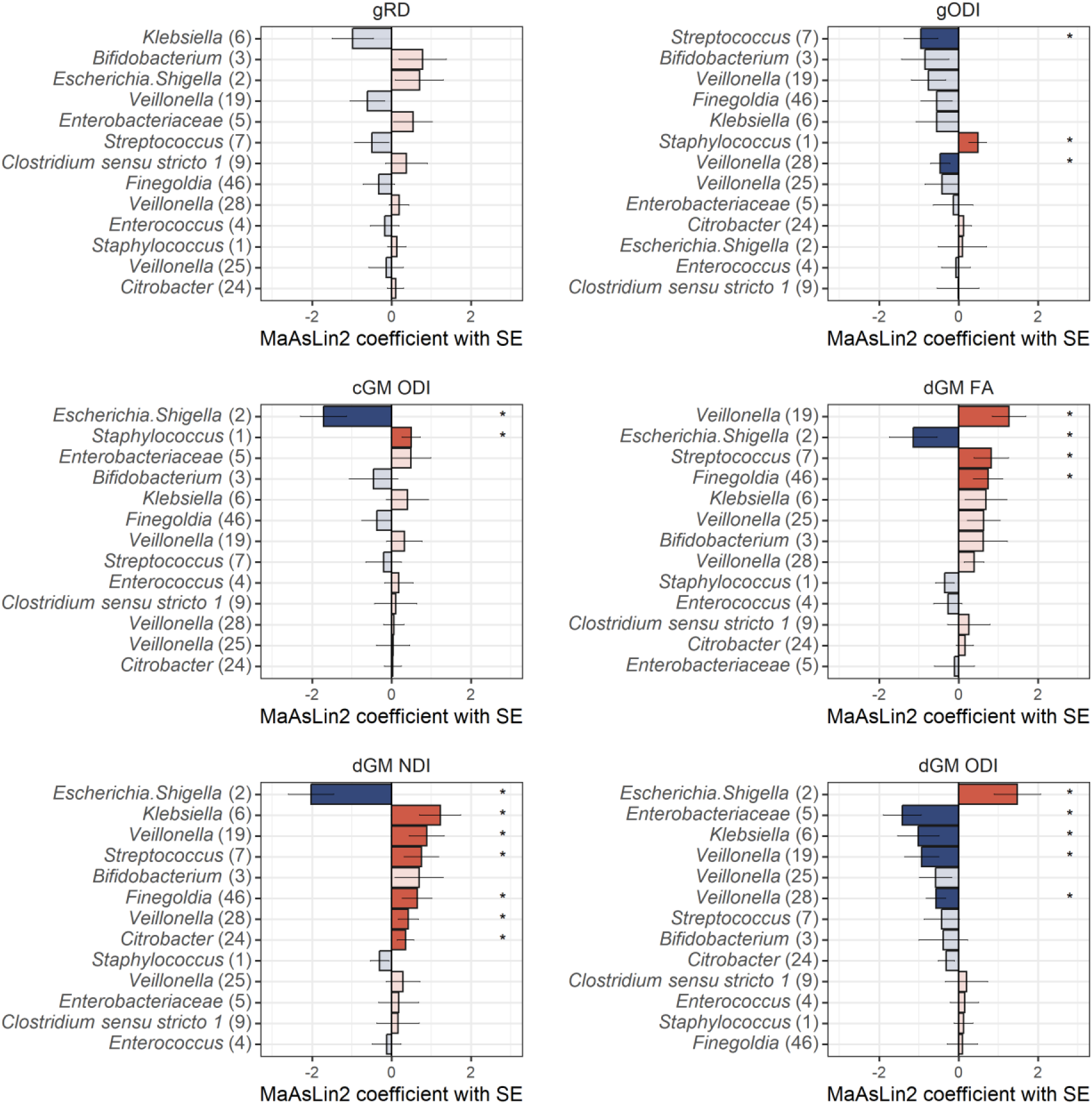
Taxa-level analyses correlating brain microstructural features with the relative abundances of ASVs. Analyses were conducted using MaAsLin2, testing differences in ASVs present with at least 1% of abundance in at least 10% of samples (n=13 ASVs). ASVs are ordered by the strength of association with each brain imaging feature. Lengths of the bars correspond with the MaAsLin2-model coefficient, which relates to the strength of the association. Error bars indicate the standard error (SE) of the model coefficient. Full colour bars and asterisks (*) indicate Benjamini-Hochberg (BH)-method-adjusted p-value < 0.25. Red indicates positive and blue negative associations. ASV = amplicon sequence variant; MaAsLin = Microbiome Multivariate Association with Linear Models; FA = fractional anisotropy; RD = radial diffusivity; NDI = neurite density index; ODI = orientation dispersion index; cGM = cortical grey matter; dGM = deep grey matter, g = general factor.

These analyses revealed further relationships between brain MRI features and bacterial taxa beyond the main drivers of the community composition variance. Notably, different *Veillonella* ASVs positively correlated with dGM FA/NDI (q range 0.018 – 0.209) and negatively with ODI (q range 0.083 – 0.115).

Analyses using the shotgun data showed similar bacteria-brain associations (Table S10). Importantly, these replicated the *E.coli*, *Klebsiella* spp. (*oxytoca, michiganesis, pneumoniae*) and *Veillonella parvula* correlations with dGM microstructure, and *E. coli* correlations with cortical ODI. Interestingly, using these data, *Bifidobacterium breve* positively correlated with FA in the dGM.

### BACTERIAL FUNCTIONAL CAPACITY AND BRAIN MICROSTRUCTURE

To probe potential functional implications of the bacteria-brain relationships, we calculated gut-brain modules^47^ (GBMs) from the shotgun data. Among the most abundant GBMs were several related to excitotoxic pathways including glutamate and quinolinic acid metabolism (Figure 6A).

**Figure 6.**
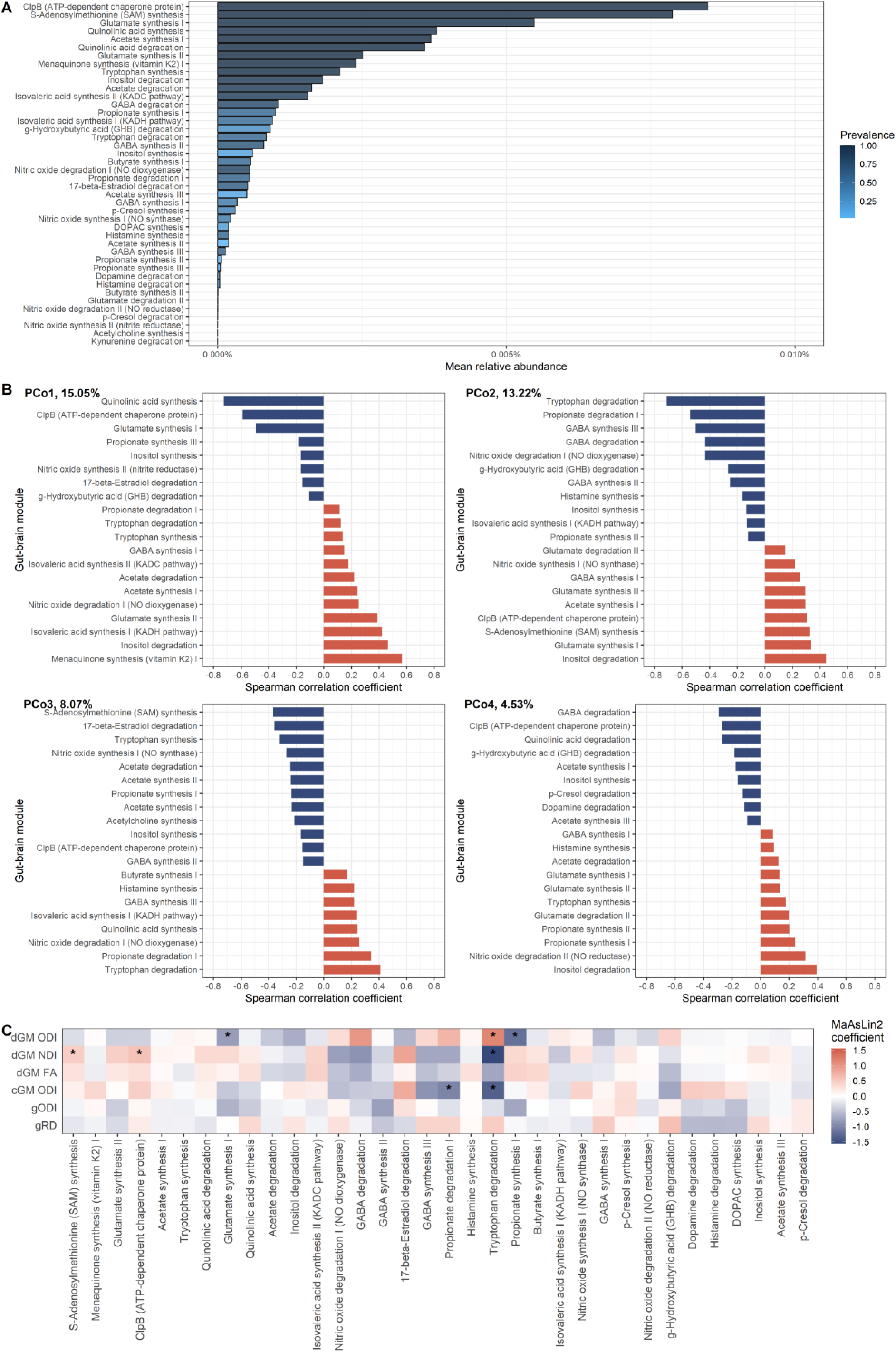
Gut-brain modules in association with brain microstructure in preterm infants. (A) Mean relative abundance of the gut-brain modules reflecting functional potential of the metagenome; bars are coloured by the prevalence of the modules. 42 out of 56 GBMs were detected in the microbiome-MRI matching dataset; all are present in at least two samples. (B) Gut-brain module correlations with the first four orthogonal principal coordinates (PCo) calculated from 16S rRNA beta diversity data, showing the top 20 strongest correlations for each PCo. The % refers to the variance explained by each of the PCo. Red indicates positive and blue negative correlations between the PCo-s and gut-brain modules. (C) Gut-brain modules in correlation with brain microstructural features. Analyses were conducted using MaAsLin2, testing differences in gut-brain modules present in at least 10% of samples (n=34 modules). Modules are ordered (left to right) by the prevalence in the dataset. Colour corresponds to MaAsLin2-model coefficient, which relates to the strength of the association, with blue indicating negative and red positive correlations. Asterisks (*) indicate Benjamini-Hochberg (BH)-method-adjusted p-value < 0.25. FA = fractional anisotropy; RD = radial diffusivity; NDI = neurite density index; ODI = orientation dispersion index; cGM = cortical grey matter; dGM = deep grey matter, g = general factor. See Table S11 for MaAsLin2 results and Figure S4 for species contribution to GBMs.

PCo1 correlated the strongest with modules related to quinolinic acid and menaquinone synthesis; PCo2 correlated with modules related to degradation of tryptophan, and inositol; PCo3 correlated with modules for the synthesis of S-adenosylmethionine (SAM) and degradation of tryptophan; and PCo4 correlated with GABA and inositol degradation (Figure 6B).

We then studied the relationships between GBM abundances and those MRI markers that were identified as significantly associated with the gut microbiota PCo-s or alpha diversity using MaAsLin2. This revealed the strongest associations between modules related to the capacity of propionate and tryptophan metabolism and NODDI measures in the deep and cortical grey matter (Figure 6C, Table S11). dGM microstructure additionally associated with modules related to caseinolytic peptidase B (ClpB), SAM and glutamate synthesis – three most abundant GBMs identified.

Lastly, to understand which bacteria could contribute to the brain-associated GBMs, we studied the correlations between species and module abundances as well as the species-stratified module abundances. In line with PCo-GBM correlations (Figure 6C), we found correlational evidence that *E. coli* is the strongest contributor to the modules related to the capacity of tryptophan and propionate degradation, and *Klebsiella* spp. have high contributions to the capacity of propionate synthesis (Figure S4). *Bifidobacterium* spp. were among the main contributors to the most abundant GBMs.

## DISCUSSION

We characterised the gut microbiome of preterm infants using 16S rRNA gene and shotgun metagenomic sequencing, and determined the most influential clinical drivers of the preterm microbiota during NICU care. For the first time, we integrated metagenome data with multimodal brain MRI to uncover associations between microbiota community composition, diversity and functional capacity, and the EoP.

### NEONATAL MICROBIOTA DEVELOPMENT

Consistent with previous reports of a dynamic development of the gut microbiota over the neonatal period in preterm infants^30,48^, there was a substantial shift in microbiota diversity and community composition between preterm birth and hospital discharge. Shortly after birth, the microbiota of the majority of infants was dominated by the facultative anaerobe S*taphylococcus*. By the time of NICU discharge, the microbiota diversity had increased and infants had gut microbiota profiles with high relative abundances of either *Bifidobacterium* or *Enterobacteriaceae*, mainly *Klebsiella* spp.; community compositions high in *Escherichia/Shigella* or *Enterococcus* were also prevalent.

### DIFFERENCES IN THE MECONIUM MICROBIOTA OF TERM AND PRETERM INFANTS

There were small differences in bacterial community composition between term and preterm meconium with higher abundance of *Escherichia/Shigella* and *Streptococcus* in the preterm group – a profile that has been reported previously^49^. The higher microbiota richness in preterm meconium might reflect the slightly higher postnatal age at sampling in the preterm group due to delayed passage of meconium in preterm infants, and is consistent with a decrease in alpha diversity in the first days of life due to environmental filtering before a gradual increase coinciding with actual colonisation^30,50^. An important technical consideration for interpreting meconium microbiota data is that only a small proportion of term infant meconium samples had sufficient bacterial biomass for sequencing. Yet, it has been reported previously that meconium has low bacterial DNA^51,52^, with relatively higher detection in preterm infants^51^. Even among the preterm group, only a minority of meconium samples had sufficient DNA yield for shotgun sequencing. Thus, the profiles may not be representative of the majority of meconium samples; this could explain some of the differences observed in the results with 16S-based vs shotgun sequencing.

### DRIVERS OF THE PRETERM GUT MICROBIOTA AT BIRTH AND AT NICU DISCHARGE

In line with studies in term infants^33,34^, delivery mode had the strongest correlation with the bacterial community composition shortly after birth. There is variability in the preterm literature about the impact of mode of delivery^22^; these data provide additional information from a new cohort. At TP2, delivery mode was not associated with microbiota composition, which could reflect either “recovery” of gut microbiota profiles ^33^, or an overshadowing by prematurity-related co-exposures. Indeed, low birth GA was the strongest covariate correlating with bacterial community composition at TP2, which suggests there is an allostatic load of prematurity-related co-exposures shaping the preterm microbiota. Extremely preterm infants had higher abundance of *Veillonella* – a signature taxa in the 4-month-old term infant microbiota, associated with reduced oxygen concentration and utilisation of lactic acid^32^. Given that low GA also associated negatively with the abundance of *Staphylococcus*, these findings suggest a “younger-looking” microbiota at the time of NICU discharge in infants born at a higher GA. Infant sex and antibiotic exposure also contributed to microbiota composition at TP2. Only a few studies have investigated the role of sex in microbiota development, yielding mixed results^30,44,53,54^. We found the strongest evidence for higher abundance of *Streptococcus* in male babies. Sex differences in this context are interesting because preterm boys have a higher risk of major morbidities than preterm girls^45^. In line with previous studies^36^, we report higher abundance of *Klebsiella* in infants exposed to antibiotics during their NICU stay; *Enterobacteriaceae*, including *Klebsiella* spp. correlate with increased antimicrobial resistance genes in antibiotic-exposed preterm infants^19,35^. Similarly supporting previous findings^36^, exposure to antibiotics rather than length of exposure associated with microbiota composition in preterm infants, suggesting an exquisite sensitivity of the preterm microbiota to antimicrobial treatment.

### MICROBIOTA-BRAIN INTERACTIONS: RELEVANCE TO ENCEPHALOPATHY OF PREMATURITY

We took two complementary approaches to study correlations between the microbiome and brain structure: we applied dimensionality reduction^55^ to construct latent variables capturing the main variance of bacterial composition, followed by post-hoc MaAsLin2 analyses.

Results from both methods suggested the strongest correlations between the relative abundances of *Escherichia/Shigella* (*E. coli* from shotgun sequencing) and dGM and cortical microstructure, particularly measures derived from NODDI, and between *Enterobacteriaceae* (*Klebsiella* spp. using shotgun sequencing) and dGM microstructure. This suggests that gut microbiota associates with cellular and dendritic morphology as previously demonstrated in rodent models of microbiota disruptions^56^. Morphological changes in dGM nuclei are commonly observed in preterm infants^57–59^; these associate with reduced microstructural integrity in the white matter and poorer neurodevelopmental outcomes^5^. In addition, preterm birth associates with alterations in cortical microstructure and morphology at TEA^60^. *Escherichia/Shigella* and *Klebsiella* have been linked with neurodevelopment previously, primarily with better and worse outcomes, respectively^24,26,28,61^. Thus, the current bacteria-brain findings are intriguing, given the age- and birth GA-associations for the NODDI parameters (Table S7). For example, dGM NDI is positively and ODI negatively associated with GA at scan, suggesting that decreased abundance of *Escherichia* and increased abundance of *Klebsiella* spp. associate with microstructural markers related to more mature dGM microstructure. However, the functional/behavioural implications of microstructural changes in the neonatal grey matter are not yet established, leaving uncertainty in the assignment of positive/negative valence to the bacteria-brain relationships. Nevertheless, these findings contribute to the literature highlighting the importance of these prevalent bacteria in the microstructural development of deep and cortical grey matter in preterm infants.

Post-hoc analyses also revealed bacteria-brain relationships that were not captured as the main drivers of the first four beta diversity PCo-s. Specifically, *Veillonella parvula*, which associated with degree of prematurity, correlated with dGM microstructural parameters, following the same direction of effect as GA at scan (Table S7). *Veillonella* has been associated with neurobehavioural outcomes, including motor and temperament development^13^. *Veillonella* could play different roles in brain function and behaviour at different developmental phases, and it remains to be established to what extent these relationships may be mediated by brain microstructure around the time of birth.

Calculation of GBMs allowed functional interpretation of the bacteria-brain relationships. In particular, the PCo indicated by the abundances of *Escherichia* and *Enterobacteriaceae/Klebsiella oxytoca*, correlated the strongest with the capacity for tryptophan degradation. Indeed, MaAsLin2 analyses showed that this module’s abundance correlated with the microstructure in dGM and cortex. Based on species-stratified gene annotations and species-module correlations, *E. coli* was the most substantial contributor to this functional property of the microbiome. Tryptophan metabolism has been suggested as one of the key gut-brain communication mechanisms in preclinical and human observational studies^10^. *E. coli* degrades tryptophan into indole, which regulates gut epithelial cell function and immune response^62^, as well as central nervous system inflammation via astrocytes^63^. Thus, our results suggest that *E. coli* may interact with brain microstructural development via tryptophan metabolism, but future mechanistic work and studies incorporating metabolome analysis are needed to further study this relationship.

Synthesis and degradation of the short-chain fatty acid (SCFA) propionate correlated with ODI in dGM and cortex, respectively. The strongest contributors to these modules were *Klebsiella* spp. And *E. coli*, respectively. Propionate, acetate and butyrate are among the most abundant SCFAs in the human body. SCFAs have wide-ranging functions^64^, including (neuro)immune modulation, and propionate has been demonstrated to impact the blood-brain-barrier^65^. Future studies, including metabolomics, are required to validate the current findings, and to identify to what extent bacterial-derived propionate directly interacts with the brain.

*Bifidobacterium* is the predominant bacterium in vaginally delivered breastfed infants during the first year of life, and several studies have identified positive correlations between *Bifidobacterium* abundance and neurobehavioural outcomes^13^, including in preterm infants^25^. It is sometimes used as a probiotic to prevent NEC in preterm infants, though clinical efficacy is uncertain and we are not aware of studies of its impact, if any, on neurodevelopment^66^. We found some suggestive evidence that the primarily *Bifidobacterium*-driven PCo1 correlates with total brain and white matter volume and with relative dGM volume, reflecting improved brain growth in association with higher abundance of *Bifidobacterium*. We also found that *Bifidobacterium* spp. were one of the main contributors to the three most abundant GBMs in this preterm dataset, which significantly correlated with dGM microstructure: ClpB, SAM and glutamate synthesis. This may suggest that *Bifidobacterium* involvement in metabolic pathways may be important for brain structural development. However, these potential relationships between *Bifidobacterium* and brain structure need to be replicated in an independent cohort. A recent Cochrane review^66^ concluded that further large, high-quality trials are needed to inform clinical practice about probiotic use for the prevention of NEC in preterm infants. Our data indicate that assessment of neurodevelopment should be incorporated into future studies of safety and efficacy of probiotics in preterm infants.

### STRENGTHS AND LIMITATIONS

The main strength and uniqueness of this study is the linked microbiome-MRI dataset. There is a scarcity of metagenomics data alongside multi-modal neuroimaging, particularly in the neonatal population, making this a valuable contribution to the microbiota-neuroimaging field. We evaluated infants born at <32 weeks’ gestation, who are at an especially high risk of adverse neurocognitive outcomes. The clinical profile of the cohort, the absence of major parenchymal lesions, and the similarities of microbiota community composition with other studies, suggest that it is representative of the majority of survivors of neonatal intensive care.

Low biomass samples, such as those collected from neonates, are at an increased risk for biases due to possible bacterial DNA contamination from processing reagents and environment^67^. We included control samples, which enabled to investigate the environmental background and remove contaminant taxa from the dataset. Furthermore, the inclusion of shotgun metagenomic sequencing alongside 16S enabled us to investigate within-study replicability and probe functional capacity.

The study has some limitations. Whilst the sample size was larger than that of previous work integrating microbiota with neuroimaging in infancy and childhood^28,55,68–72^, it is still a limitation given the high inter-individual variation both in brain microstructural as well as in the microbiota development in preterm infants. Small samples and high heterogeneity coupled with the multidimensional nature of both microbiota sequencing and neuroimaging datasets and high analytic freedom are the main limitations of microbiota-neuroimaging studies, leading to reduced power and variability in the results. To reduce dimensionality, we focussed on whole-brain measures capturing EoP. However, this may have hindered the detection of more specific brain regions associated with microbiota; future studies with larger sample sizes are needed to investigate the regional-specificity of the observed global effects. PCo analysis and calculation of GBMs from the microbiome data allowed for a principled way of data reduction. There is rapid development of the gut microbiota in the neonatal period, thus, the wide age range of sampling could have introduced noise in the microbiota-brain relationships; to mitigate this, we always adjusted for age at microbiota sampling. The sample size was not sufficiently powered for age- or sex-stratified analyses.

Whilst shotgun metagenomic sequencing allows evaluation of the functional capacity of the microbiome, future studies including metabolomics measurements are needed to confirm whether gene abundances equate to differences in metabolite levels. Additionally, our interpretation of bacterial species’ contributions to GBMs was based on species-GBM abundance correlations and species-stratified GBM annotations, thus, future mechanistic work using *in vitro* or animal models are needed to confirm the validity of these findings.

Finally, the microbiota-brain association analysis was cross-sectional with the timepoints chosen to capture EoP and the allostatic load of prematurity on the gut microbiome. However, this limits causal inference: the relationships observed could reflect a separate process causally linked to the development of both the gut microbiome and brain structure. Nevertheless, preclinical data shows that colonisation of germ-free mice with preterm microbial communities associates with poor growth, systemic and neuroinflammation, delayed neuronal development and myelination, disrupted brain microstructural connectivity, and behavioural deficits^73,74^, supporting a causal relationship. Future studies with longitudinal microbiome sampling over the NICU period are needed to clarify the critical time window for the strongest influence of the gut microbiome on brain microstructural development. This is important to identify the optimal time for microbiome modification-based therapies for brain health following preterm birth.

In conclusion, the results contribute to understanding microbiota-brain associations following preterm birth and suggest that microbiota modification is a potential new avenue for neuroprotection during neonatal intensive care.

## Data Availability

All raw 16S and shotgun data and their derivates used for analysis alongside with participant clinical data, sample metadata, and neuroimaging data derivates used in this work are deposited in Edinburgh DataVault (https://doi.org/10.7488/e65499db-2263-4d3c-9335-55ae6d49af2b). Requests for access will be considered under the study's Data Access and Collaboration policy and governance process (https://www.ed.ac.uk/centre-reproductive-health/tebc/about-tebc/for-researchers/data-access-collaboration). The microbiome and participant metadata reported in this study cannot be deposited in a public repository due to them containing information that could compromise participant consent. Requests for raw neuroimaging data will similarly be considered under the study's Data Access and Collaboration policy and governance process: https://www.ed.ac.uk/centre-reproductive-health/tebc/about-tebc/for-researchers/data-access-collaboration.
Code used for the data analysis in this paper is available here: https://git.ecdf.ed.ac.uk/jbrl/neonatal-microbiota-and-brain-dysmaturation.

## AUTHOR CONTRIBUTIONS

KV, DB and JPB conceived and designed the experiments. GS, DQS and AC recruited the participants. GS, DQS, AC and KV supervised MRI acquisition and collected clinical data. PLP and KV were responsible for the execution and quality control of the laboratory work. MBC and KV performed MRI data quality control and processing. AJQ was responsible for clinical evaluation of MR images. MJT and MEB designed MRI acquisition protocols. KV and JB performed bioinformatic processing. KV analysed the data. JB, GJVB and MLO assisted with data analysis. KV, DB and JPB wrote the paper. All authors significantly contributed to interpreting the results, critically revised the manuscript for important intellectual content and approved the final manuscript.

## ACKNOWLEDGEMENTS

This research was funded in whole, or in part, by the Wellcome [No. 108890/Z/15/Z]. For the purpose of open access, the author has applied a CC BY public copyright licence to any Author Accepted Manuscript version arising from this submission. This work was supported by Theirworld (www.theirworld.org) and was carried out in the Medical Research Council Centre for Reproductive Health, which was funded by Medical Research Council Centre Grant (MRC G1002033). Participants were scanned in the University of Edinburgh Imaging Research MRI Facility at the Royal Infirmary of Edinburgh, which was established with funding from The Wellcome Trust, Dunhill Medical Trust, Edinburgh and Lothians Research Foundation, Theirworld, The Muir Maxwell Trust and other sources. KV was funded by the Wellcome Translational Neuroscience PhD Programme at the University of Edinburgh (108890/Z/15/Z). MJT was funded by the NHS Lothian Research and Development Office. The authors are grateful to the families who consented to take part in the study and to all the University’s imaging research staff for providing the infant scanning. We would also like to acknowledge Edinburgh Genomics for executing the 16S rRNA gene sequencing and Heleen de Weerd for the bioinformatic processing of the shotgun data; shotgun metagenomic sequencing was performed at Novogene.

## STAR METHODS

### RESOURCE AVAILABILITY

#### Lead contact

Further information and requests for resources and reagents should be directed to and will be fulfilled by the Lead Contact, James P Boarman.

#### Materials availability

This study did not generate new unique reagents.

#### Data and code availability

All raw 16S and shotgun data and their derivates used for analysis alongside with participant clinical data, sample metadata, and neuroimaging data derivates used in this work are deposited in Edinburgh DataVault^75^ (https://doi.org/10.7488/e65499db-2263-4d3c-9335-55ae6d49af2b). Requests for access will be considered under the study’s Data Access and Collaboration policy and governance process (https://www.ed.ac.uk/centre-reproductive-health/tebc/about-tebc/for-researchers/data-access-collaboration). The microbiome and participant metadata reported in this study cannot be deposited in a public repository due to them containing information that could compromise participant consent. Requests for raw neuroimaging data will similarly be considered under the study’s Data Access and Collaboration policy and governance process: https://www.ed.ac.uk/centre-reproductive-health/tebc/about-tebc/for-researchers/data-access-collaboration.

Code used for the data analysis in this paper is available here: https://git.ecdf.ed.ac.uk/jbrl/neonatal-microbiota-and-brain-dysmaturation.

### EXPERIMENTAL MODEL AND STUDY PARTICIPANT DETAILS

#### Study participant details

Participants were preterm infants (GA at birth <33 weeks) and term-born controls recruited as part of a longitudinal cohort study designed to investigate the effects of preterm birth on brain structure and long-term outcome^29^. Recruitment, sampling and MRI acquisition were at the Royal Infirmary of Edinburgh, UK, between 2016-2021. The study was conducted according to the principles of the Declaration of Helsinki, and ethical approval was obtained from the UK National Research Ethics Service (South East Scotland Research Ethic Committee 16/SS/0154). Parents provided written informed consent.

Exclusion criteria were death during neonatal period, major congenital malformations, chromosomal abnormalities, congenital infection; infants with overt parenchymal lesions (cystic periventricular leukomalacia, haemorrhagic parenchymal infarction), post-haemorrhagic ventricular dilatation, or contra-indications to MRI were excluded from MRI analyses. Term-born infants who required admission to the NICU were also excluded.

All infants were cared for in the Neonatal unit of the Simpson Centre for Reproductive Health, Royal Infirmary of Edinburgh, with standardised feeding, antibacterial and antifungal guidelines. Preterm infants admitted to the NICU in the Simpson Centre for Reproductive Health are not routinely administered any pro- or prebiotic supplements. Clinical data was collected from antenatal and neonatal electronic patient records.

Clinical variable definitions Incidence of neonatal sepsis (early or late onset) was defined as detection of a bacterial pathogen from blood culture, or physician decision to treat with antibiotics for ≥ 5 days in the context of growth of coagulase negative *Staphylococcus* from blood or a negative culture but raised inflammatory markers in blood. Necrotising enterocolitis (NEC) was defined as stages II or III according to the modified Bell’s staging for NEC which required medical treatment for ≥ 7 days or surgical treatment, respectively^76^. Bronchopulmonary dysplasia (BPD) was defined as the requirement for supplemental oxygen or respiratory support at 36 weeks gestational age. Retinopathy of prematurity (ROP) was defined as requiring treatment with laser therapy or anti-VEGF. Birthweight z-scores were calculated according to International Fetal and Newborn Growth Consortium for the 21st Century (INTERGROWTH-21st) standards for preterm infants^77^.

Antibiotic exposure was assessed by three composite variables: (i) exposure to antibiotics during the first three days of life, (ii) exposure to antibiotics at any other time during the NNU stay, and (iii) proportion (%) of antibiotic exposure days during NNU stay (total number of antibiotic treatment days was divided by the number of days in NNU). The antibiotic treatment for all preterm infants with suspected and confirmed neonatal sepsis conformed to the following principles: babies up to 72 h of age were commenced on benzylpenicillin and gentamicin, babies > 72 h of age were commenced on piperacillin/tazobactam and vancomycin. To reduce unnecessary exposure to antibiotics, treatment was stopped after 48 h if blood cultures were negative and the clinician had a low suspicion about infection. Some infants in the cohort were also treated with azithromycin, cefotaxime, co-amoxiclav, flucloxacillin, linezolid, meropenem and metronidazole according to symptoms and diagnostic results.

Daily nutritional intake for preterm infants was collected from birth until discharge. Each day was categorised as consisting of exclusive maternal breast milk feeds, exclusive formula milk feeds, exclusive donor expressed milk feeds, or any combination of these feeding types. Data was available as the sum of each feeding type over the entire duration of NNU stay. As previously^78,79^, exclusive breast milk exposure was defined as the % of inpatient days that infants received exclusive breast milk feeds, which included both maternal and/or donor breast milk. Infants were categorised into two groups based on breast milk exposure: high breast milk exposure was defined as exclusive breast milk feeds for ≥ 75% of inpatient days and low breast milk exposure was defined as exclusive breast milk feeds for < 75% of inpatient days.

### METHOD DETAILS

#### Faecal sample collection and processing

Faecal material was collected from dirty diapers by parents, NICU staff or research team. The samples were frozen at −20°C directly after sample collection prior to transfer to a −80°C freezer in the Queens Medical Research Institute (QMRI, University of Edinburgh) until further analyses; no preservation buffers were used. Faecal material was collected from the first bowel movement (meconium; TP1) from term and preterm infants, and a second faecal sample was collected from preterm infants prior to discharge from the NICU (pre-discharge sample; TP2), which was around TEA. When preterm infants were transferred to another NICU prior discharge, the second sample was collected prior to transfer. A total of 143 meconium and 107 pre-discharge samples were collected during the study period; 44 preterm infants had both samples obtained.

#### DNA isolation

The bacterial DNA from faecal samples was extracted at the QMRI as previously described^33,80^ involving phenol/bead beating in combination with the Mag Mini DNA Isolation Kit (LGC genomics, Germany). Samples were thawed on ice for as little time as possible to obtain one 10 μl inoculation loop of raw faeces which was added to a 2 ml screwcap tubes containing a mixture of 150 μl lysis buffer (Mag Mini DNA Isolation Kit, LGC genomics, Germany), 0.1 mm zirconium beads (BioSpec products, USA) in 650 μl lysis buffer, and 500 μl of phenol saturated with Tris-HCl (pH 8.0; BioSpec products, USA). The samples were mechanically disrupted twice for 2 minutes at 2100 oscillations/minute using a bead beater (BioSpec products, USA). The samples were then centrifuged for 10 minutes at 5000 rpm at room temperature. Then, the aqueous phase was added to 1300 μl of binding buffer (Mag Mini DNA Isolation Kit) with 10 μl magnetic beads (LGC genomics, Germany) in a sterile 1.5 ml Eppendorf tube and incubated for 30 minutes at room temperature on a thermos shaker (Hettich lab technologies, USA) to allow DNA binding. Subsequently, the supernatant was discarded. The magnetic beads were washed twice with wash buffer 1 (Mag Mini DNA Isolation Kit,), once with wash buffer 2 (Mag Mini DNA Isolation Kit), and air-dried for 15 minutes at 55°C. DNA was eluted in 50 μl elution buffer. Adaptions in the standard DNA isolation procedure were applied for timepoint 1 samples due to low DNA yield and interference with extraction protocol. Thus, the following adaptions were applied: (i) two inoculation loops of faeces were used as input material; (ii) lysis buffer volume was increased to 200 μl; (iii) addition of six 2 mm glass beads (Scientific Laboratory Supplies, UK) for more efficient mechanical sample disruption; (iv) to improve water phase separation, the initial centrifugation was increased to 15 minutes and an additional centrifugation for 5 minutes was performed for some samples to improve separation of the aqueous phase; however, when there was little separation of the aqueous layer, more volume from the other layers was included in the next extraction steps; (v) the washing steps were performed with 400 μl of the buffers; (vi) the DNA was eluted in a final volume of 35 μl to increase final DNA concentration.

To avoid potential cross-contamination from high-abundant to low-abundant samples, DNA from meconium and pre-discharge faecal samples was isolated on separate days. Each extraction was accompanied by negative (200 μl of lysis buffer) and positive controls (ZymoBIOMICS Microbial Community Standard [Zymo Research, USA] and/or a convenience saliva sample).

The amount of extracted bacterial DNA was determined by quantitative polymerase chain reaction (qPCR) as previously^80,81^ with universal primers and probes targeting the 16S-rRNA gene (forward: 5′-CGAAAGCGTGGGGAGCAAA-3′, reverse: 5′-GTTCGTACTCCCCAGGCGG-3′, TAMRA probe: 6FAM-ATTAGATACCCTGGTAGTCCA-MGB; Life Technologies, USA).

#### 16S rRNA gene sequencing

Samples that yielded DNA concentration of >0.18 pg/μl were considered for 16S rRNA gene sequencing (Figure S1).

V4 hypervariable region of the 16S rRNA gene was amplified as previously^82^: amplicon libraries were generated by PCR using barcoded primers (515F [5’-GTGCCAGCAGCCGCGGTAA-3’[ and 806R [5’-GGACTACCAGG-GTATCTAAT-3’]^83^), using 5μl of DNA as template. Two mock DNA communities (see below) and a non-template control were included in each MiSeq PCR plate and amplified alongside the samples and isolation positive and negative controls.

The DNA mock communities used alongside samples for the amplification of V4 hypervariable region of the 16S rRNA gene were: equimolarly pooled bacterial DNA from eleven species (*Bacteroides fragilis*, *Haemophilus influenzae*, *S. pneumoniae*, *Streptococcus pyogenes*, *Klebsiella oxytoca*, *Klebsiella pneumoniae*, *haemolytic Streptococcus group A*, *Pseudomonas aeruginosa*, *Staphylococcus epidermidis*, *Staphylococcus aureus* and *Moraxella catarrhalis*); and the ZymoBIOMICS Microbial Community DNA Standard (Zymo Research, USA).

The amplified DNA concentration was quantified using Quant-iT™ PicoGreen® dsDNA Assay Kit (Thermo Fisher Scientific, USA) and visualised on gel electrophoresis to ensure successful amplification. The amplicons were pooled at equimolar amounts and purified using a combination of agarose gel purification (GeneJET Gel Extraction and DNA Cleanup Micro Kit) and purification by AMPure XP magnetic beads (Thermo Fisher Scientific, MA, USA).

As previously^82^, 16S rRNA gene sequencing was performed using the MiSeq Reagent Kit v2 on the Illumina MiSeq platform (Illumina, USA). Sequencing was performed by Edinburgh Genomics (University of Edinburgh, UK) on a total of 191 samples, 23 negative and 18 positive controls in one run.

#### Bioinformatic processing and quality control of 16S rRNA sequences

16S rRNA gene sequencing data processing was performed in R (version 4.2.1)^84^ as previously described^85^. Paired-end raw reads were filtered and trimmed (maxEE = 2; truncLen = 200/150 bp for forward and reverse reads, respectively), merged, denoised, chimera filtered and binned into ASVs using the DADA2 (version 1.16.0) in R^86^. Taxonomy was assigned using the DADA2 implementation of the naïve Bayesian classifier using the Silva v138.2 reference database^87^. Species-level annotations were added using the *addSpecies()* function. ASVs not assigned to the kingdom Bacteria or assigned to the family Mitochondria or class Chloroplast were removed.

Contamination was assessed using *decontam* package in R^88^ (*isContaminant* function, “combined” method, default parameters), combined with manual inspection of putative contaminating ASVs. Using *decontam*, DNA extraction blanks were used as negative controls and values from 16S qPCR were used for the measure of DNA concentrations. To ensure the accuracy of the method, these contaminant ASVs (n=72) were carefully inspected by plotting the 16s qPCR DNA concentration data against the relative abundance. Second, in order to exclude ultra-rare taxa from the final dataset, the ASV table was filtered by removing ASVs that were identified at a relative abundance of <0.1% and present in less than two samples^89^. Thereafter, the remaining list of ASVs were cross-matched to those taxa identified as contaminants by Salter et al^90^. These ASVs were manually inspected by plotting the 16s qPCR DNA concentration data against the relative abundance per isolation batch. Contaminant species were defined as ASVs abundant only in the lowest density samples in each isolation and/or only in isolation blanks. These additional contaminant ASVs (n=27) were then additionally removed from the raw ASV list after which filtering of the ultra-rare taxa was repeated. After excluding contaminating and ultra-rare taxa, the number of remaining reads per sample was investigated. Samples that had a final read count of less than 5000 (n=8) were excluded from the final dataset; these 8 samples also had >70% of the reads removed during the decontamination process, suggesting the remaining reads may not reliably represent the community composition. Duplicate samples (n=4) were also excluded from the final dataset. These quality control steps removed a mean of 3.75% of the raw reads from the dataset. Following data quality control, the 16S-based sequencing produced a mean of 35416 reads (range 7534 – 109456) per sample. The ASV table contained 174 ASVs.

In the final analytic sample, all participants with TP2 samples and 70/136 (51.5%) TP1 samples were included (Figure S1). Included participants with TP1 samples were more likely obtained from preterm participants and were thus collected at a later gestational and postnatal age.

#### Metagenomic shotgun sequencing and bioinformatic processing

Samples with 16S qPCR concentration >0.8ng/μl (n=121) alongside three saliva positive controls and eight isolation negative controls were considered for shotgun metagenomic sequencing at Novogene facility (Novogene Co., Ltd, Cambridge, UK). Sequencing was performed on the NovaSeq 6000 platform (Illumina) with a read length of 150-bp paired-end reads producing 9G raw data per sample. Shotgun sequencing failed for all negative controls, indicating absence or very low abundance of biological material, and one biological sample from TP1.

Data pre-processing and annotation was performed by Edinburgh Genomics. Whole metagenome shotgun sequencing produced a mean of 37410649 (range 2512236 – 81101422) raw reads per sample. The raw reads were cleaned using cutadapt (v3.5)^91^. Adapters were removed, reads were cut when the quality dropped below 30, and reads shorter than 50 bases were removed. Reads belonging to the host were removed by bowtie2 (v2.4.1)^92,93^ using Homo Sapiens (GRCh38) as a reference. Files sequenced on multiple lanes but belonging to the same sample were merged into single forward and reverse files.

Taxonomic profiling was performed using MetaPhlAn (v3.1)^94^ with the standard database (mpa_v31_CHOCOPhlAn_201901). Functional profiling was performed using HUMAnN (v3)^94^ with the default chocophlan (chocophlan.v201901_v31) and the uniref90 databases (uniref90_annotated_v201901b). As MetaPhlAn and HUMAnN do not use paired information of reads, all reads of a sample were merged into a single file and used for taxonomical/functional assignment. As shotgun metagenomic sequencing was performed on higher density samples, the relative contribution of potentially contaminant taxa is smaller, thus, no further quality controls/decontamination on species/functional level were performed. Functional gene families data was grouped to KOs using the *humann_regroup_table* function, both for community-level totals and species-stratified gene families. The KO abundance data table was total-sum-scaled (TSS) to relative abundances using the *humann_renorm_table* function; the unmapped and ungrouped reads were taken into account for TSS-normalisation, but excluded from downstream statistical analyses. From the normalised KO table we computed GMMs^96^ and GBMs^47^ using the omixer-rpmR library^95^ using the default settings for both community and taxon-stratified levels. Shotgun sequencing dataset contained a total of 223 species, 94 GMMs, and 43 GBMs.

#### Magnetic resonance imaging data acquisition

Infants were scanned at TEA at the Edinburgh Imaging Facility, Royal Infirmary of Edinburgh, University of Edinburgh, UK, using a Siemens MAGNETOM Prisma 3T MRI clinical scanner (Siemens Healthcare, Erlangen, Germany) and a 16-channel phased-array paediatric head coil in natural sleep as previously described^98^. Each acquisition was inspected contemporaneously for motion artefact and repeated if there had been movement but the baby was still sleeping; diffusion MRI acquisitions were repeated if signal loss was seen in three or more volumes.

MRI acquisition protocols are detailed in the study protocol paper^29^. The following sequences were used in this study: a T2-weighted (T2w) sampling perfection with application optimised contrasts by using flip angle evolution (SPACE) structural scan (repetition time [TR] = 3200 ms, echo time [TE] = 409 ms, acquisition plane = sagittal, voxel size = 1 mm isotropic, FOV = 128 mm, acquisition time = 2:13 min), and a multishell axial diffusion MRI (dMRI) scan. dMRI was acquired in two separate acquisitions to reduce the time needed to reacquire any data lost to motion artefacts: the first acquisition consisted of 8 baseline volumes (b = 0 s/mm2 [b0]) and 64 volumes with b = 750 s/mm^2^; the second consisted of 8 b0, 3 volumes with b = 200 s/mm^2^, 6 volumes with b = 500 s/mm^2^ and 64 volumes with b = 2500 s/mm^2^ (acquisition time = 4:29 + 5:01 min). An optimal angular coverage for the sampling scheme was applied^100^. In addition, an acquisition of 3 b0 volumes with an inverse phase encoding direction was performed (acquisition time = 0:28 min). All dMRI images were acquired using single-shot spin-echo echo planar imaging (EPI) with 2-fold simultaneous multislice and 2-fold in-plane parallel imaging acceleration and 2 mm isotropic voxels; all three diffusion acquisitions had the same parameters (TR/TE 3400/78.0 ms).

Structural images were reported by a paediatric radiologist with experience in neonatal MRI (AJQ). Images with evidence of post-haemorrhagic ventricular dilatation, cystic periventricular leukomalacia or central nervous system malformation were excluded from subsequent analysis.

#### Imaging data pre-processing

Details on dMRI processing have been previously published^98^ using MRtrix3^103^ and FMRIB Software Library (FSL)^104^. Briefly, for each subject, the two dMRI acquisitions were first concatenated and then denoised using a Marchenko-Pastur-PCA-based algorithm^105^; eddy current, head movement and EPI geometric distortions were corrected using outlier replacement and slice-to-volume registration^106,108,110,112^; bias field inhomogeneity correction was performed by calculating the bias field of the mean b0 volume and applying the correction to all the volumes^113^. The structural images were processed using the minimal processing pipeline of the developing Human Connectome Project (dHCP)^109^ to obtain the bias field corrected T2w, brain mask, tissue segmentation and the different tissue probability maps. The mean b0 EPI volume of each subject’s dMRI acquisition was co-registered to their structural T2w volume using boundary-based registration using FMRIB’s Linear Image Registration Tool (FLIRT)^114^.

From the diffusion images we calculated the DTI (FA, RD) and NODDI (NDI, ODI and ISO) maps. The DTI model was fitted in each voxel using only the b = 750 s/mm^2^ shell. NODDI maps were calculated using all shells and the recommended values of the parallel intrinsic diffusivity for neonatal brain tissues^111,115^ using the original NODDI MATLAB toolbox (http://mig.cs.ucl.ac.uk/index.php?n=Tutorial.NODDImatlab).

#### Selection of image features

##### Volumes

We calculated the volumes of the total tissue, cortical grey matter, deep grey matter, white matter, cerebellum, and the ventricles from the tissue parcellation obtained from the dHCP pipeline^109^. For cortical grey matter, deep grey matter, white matter and the cerebellum both raw and relative (i.e. normalised to total tissue volume) volumes were obtained to quantify absolute growth of the tissues as well as that relative to total brain growth, respectively.

##### Grey matter microstructure

For cortical and deep grey matter and the cerebellum, the mean DTI and NODDI metrics were calculated, using the recommended value for the parallel intrinsic diffusivity for neonatal grey matter (1.25 μm^2^/m) for NODDI map calculations^115^. For the cortex, mean gyrification index, thickness, sulcal depth, curvature and surface area as measures of cortical complexity/morphometry were also calculated^109^.

##### White matter microstructure

To capture global white matter dysmaturation, we segmented 16 major tracts and derived general factors (g-factors) for each of the DTI and NODDI metrics as described previously^98^. The only difference with the previous work was that the tracts were brought from ENA50 neonatal template space to native space via registration of the FA maps to ENA50 FA template using rigid, affine and symmetric normalization (SyN) implemented in Advanced Normalization Tools^107^. For the tracts, the NODDI metrics were calculated using the recommended values of the parallel intrinsic diffusivity for neonatal white matter (1.45 µm^2^/ms)^115^.

For contextualisation of the image features, we performed linear regression modelling for each neuroimaging feature as the outcome and GA at birth and at scan as the predictors (Table S8). Representative brain maps are provided in Figure S5.

### QUANTIFICATION AND STATISTICAL ANALYSIS

All statistical analyses were performed in R 4.2.1^84^. Visualisations were plotted using the *ggplot2*^102^, *ggpubr*^116^ and *cowplot*^117^ packages. Where necessary, distribution of variables and regression diagnostic plots were visually inspected to ensure approximate conformation to assumptions; normal distribution was additionally assessed using Shapiro-Wilk’s test. P-values from statistical tests were adjusted for multiple comparisons using the Benjamini-Hochberg method^118^ separately within each analysis type, producing q-values (see details within each subsection).

#### Beta diversity and PERMANOVA

Beta diversity was calculated as the Bray-Curtis dissimilarity^119^ matrix based on the TSS-normalised (i.e. relative abundances) ASV, species and functional GMMs tables using *vegdist* function (*vegan* package^120^). PERMANOVA, modelled by *adonis2* (*vegan* package^120^) with 1000 permutations, was used to identify differences in overall bacterial community composition. Separate PERMANOVAs were performed for all pairwise comparisons for the different sample types and timepoints; when comparing TP1 vs TP2 composition within preterm infant group we adjusted for repeated measures by constraining the permutations within participant (strata = participant ID). To assess the effects of perinatal covariates, univariable analyses were conducted for each covariate, separately for the timepoints; p-values were adjusted for multiple comparisons across the models. Results with q-value < 0.1 were considered as statistically significant and followed up using differential abundance testing.

For visualisation purposes (Figure 1A and Figure S2A), we applied hierarchical clustering (*hclust* function) on the ASV- and species-level Bray-Curtis dissimilarity matrices. Calinski-Harabasz and Silhouette width indices were used to determine whether average (ASV data) or complete (species data) linkage was optimal for the different datasets.

#### Ordination

Principal Coordinates Analysis (PCoA) was performed on the ASV-level Bray-Curtis dissimilarity matrix using the function *pcoa* (*ape* package^97^). Cailliez transformation was applied to correct for negative eigenvalues^121^. The scree plot of the eigenvalues alongside with proportion of variance explained were inspected to determine the optimum number of coordinates to extract. For taxonomic and functional interpretation of the main axes of variance (i.e. the extracted PCo-s), Spearman correlation coefficients were calculated between the PCo scores and the relative abundances of the ASVs, species, GBMs and GMMs.

#### Species contribution to gut-brain modules

To understand which bacterial species contribute to the calculated gut-brain modules^47^, we took two approaches. First, we calculated Spearman rank correlation coefficients between the relative abundances of the gut-brain modules and 25 most abundant species in the dataset. Second, we plotted the relative abundance of the gut-brain modules stratified by species. The latter approach illustrates the extent to which a species’ genes could be attributed to a gut-brain module.

#### Alpha diversity

Shannon index and the number of observed ASVs were calculated using the *estimate_richness* function (*phyloseq* package^122^). These indices were calculated based on the ASV table after removal of contaminant taxa but before filtering of the ultra-rare taxa. This full ASV table was rarefied to the minimum sequencing depth (10200 reads before filtering of ultra-rare taxa) using *rarefy_even_depth* function. Linear mixed effects modelling (*lmer* function within package *lmerTest*^99^, Satterthwaite’s method) was used to assess differences in alpha diversity indices between the sample types and timepoints, fitting participant ID as a random effect to adjust for repeated measures. For observed ASVs, the assumption of normal distribution of model residuals was violated and therefore the values were log_10_-transformed. Post-hoc analyses were conducted using the package *emmeans*^101^. P-values were adjusted for multiple comparisons for the main effects across the models for the two alpha diversity indices, and separately for the three pairwise comparisons. We then assessed the associations between perinatal covariates and alpha diversity indices in preterm infants at the two timepoints. We calculated Spearman correlation coefficients for the associations between continuous variables and alpha diversity indices; t-tests (TP2) and Wilcoxon rank-sum tests (TP1) were applied to test for the differences in alpha diversity indices between groups of infants based on categorical variables. P-values were adjusted for multiple comparisons separately for the timepoints. Covariates with q-value < 0.1 in univariable models were followed up with multivariable linear regression modelling including all covariates to adjust for the potential confounding effects between the variables.

#### Associations between gut microbiota and brain MRI features

##### Baseline linear regression models

First, the microbiota features (PCo-s and alpha diversity indices) were adjusted for GA at sample collection by fitting a linear model of each feature on GA at sample collection and retaining the residuals. We adjusted for sampling age in this manner rather than using it as a covariate in the model with the brain MRI feature as the outcome to avoid spurious correlations between GA at microbiota sampling and brain MRI features. Then, a linear regression model was performed for each residualised microbiota feature as the predictor, each MRI feature as the outcome, and GA at birth and at scan added as covariates. All values were scaled (z-transformed) before fitting the models, resulting in standardised regression coefficients. P-values were adjusted for the FDR across all models separately for volumetric and microstructural/cortical morphometric MRI features. Due to the exploratory nature of the study and given the correlated nature of the neuroimaging measures, results with q-value < 0.25 were considered as noteworthy and investigated further in sensitivity and post-hoc differential abundance analyses. FDR correction of the p-values provides a balance between type I and type II errors; 0.25 is the default threshold in MaAsLin2 and there is precedence of using this cut off in microbiota-brain/behaviour studies^70,123^.

To quantify the variance in each brain imaging metric accounted for by the microbiota PCo-s or diversity indices, the incremental R^2^ was calculated as the difference between the multiple R^2^ of each model with that from the null model including only the covariates (GA at birth and at scan) as predictors. We used analysis of variance (ANOVA) to test whether the baseline model with the microbiota feature as the predictor fit the data significantly better compared to the null model.

##### Covariate identification and adjustment (sensitivity analyses)

The clinical variables that were significantly (q-value < 0.1) associated with microbiota alpha- or beta-diversity at TP2 on ASV-level (sex, birthweight z-score, antibiotics >72h of life, and NEC) were tested for associations with brain MRI features. Two-sample t-tests were used to compare the means of normally distributed continuous features between the groups of the categorical covariates; Wilcoxon rank-sum tests were applied to compare differences in non-normally distributed MRI features; Spearman correlation analysis was used to test for significant associations between birthweight z-score and MRI features. Variables that were nominally significantly (p < 0.05) associated with at least one of the brain MRI features were added as covariates in the fully adjusted model. These variables were: birthweight z-score, sex and NEC. Only three infants in the sample were diagnosed with NEC, thus, instead of adding this variable as a covariate, a sensitivity analysis was performed excluding infants with NEC.

#### Differential abundance testing

MaAsLin2^46^ was used to identify bacterial ASV, species and functional capacity biomarkers associated with factors of interest. We applied TSS-normalisation prior to MaAsLin2 modelling. As by default in the method, we considered results with q-values < 0.25 as statistically significant.

In analyses assessing the associations between bacterial abundance and perinatal covariates, models were performed separately for the two timepoints. We tested for the effects on ASVs (16S) and species (shotgun) that were present with at least 1% of abundance (min_abundance = 0.01) in at least 5% of samples (min_prevalence = 0.05); the normalisation method within MaAsLin2 was set to “none”; all other arguments were used as by default. Baseline analyses at TP1 were adjusted for postnatal age at sample. Baseline models at TP2 were adjusted for the degree of prematurity (GA group; very vs extremely preterm) and GA at sample collection. At TP1 we didn’t adjust for GA at birth given the lack of statistically significant association in univariable PERMANOVA; at TP2 we adjusted for GA at sample collection instead of postnatal age given the high collinearity between GA at birth and postnatal age. In fully adjusted models, all clinical variables were tested simultaneously. For species, an additional model was performed for TP1 samples including labour antibiotics in the model.

To identify ASVs, species and GBMs associated with brain imaging features, MaAsLin2 was conducted for those brain MRI features that were significantly (q-value < 0.25) associated with any of the microbiota feature in the baseline models. MaAsLin2 reverses predictors and outcomes compared to PCo-based analyses, thus, to achieve alignment of the MaAsLin2 models with the initial baseline regression model, we first adjusted the brain MRI features for GA at birth and at scan by fitting a linear model of each feature on GA at birth and at MRI, retaining the residuals. These residuals were then used independently as the predictors in the MaAsLin2 models, including GA at faecal sample collection as a covariate. Due to the relatively small sample size in comparison with the number of bacterial taxa, in all analyses, we tested for effects on ASVs and species that were present with at least 1% of abundance in at least 10% of samples; other arguments were as specified above; for GBMs, the default MaAsLin2 parameters were used.

#### Reporting summary

We followed the Strengthening The Organization and Reporting of Microbiome Studies (STORMS) checklist^124^ in describing the methodology and reporting results.

## SUPPLEMENTAL ITEMS TITLES

### FIGURES

Figure S1 (relates to Table 1). Flowchart detailing the inclusion and exclusion of samples and participants in the study.

Figure S2 (relates to Figure 1). Overview of microbiome profiles in preterm neonates from shotgun metagenomic sequencing.

(A) Relative abundances of the 25 most abundant species identified across the dataset are visualised per sample. Samples are ordered based on hierarchical clustering of the Bray-Curtis dissimilarity matrix using complete linkage (see dendrogram).

(B) Mean relative abundances of the 30 most abundant gut metabolic modules in preterm infant stool at timepoint 1 (top) and 2 (bottom); bars are coloured by the prevalence of the modules at the two timepoints.

(C,D) Non-metric multidimensional scaling plot based on Bray-Curtis dissimilarity between samples at (C) species (PERMANOVA R^2^ = 3.31%, p = 0.002) (D) and gut metabolic modules (PERMANOVA R^2^ = 4.17%, p = 9.99 × 10^−4^) level; data points and ellipses are coloured by sample type. The ellipses denote the standard deviation of data points belonging to each sample type, with the centre points of the ellipses calculated using the mean of the coordinates per group. 94/103 gut-brain modules were identified in the dataset.

Sample sizes: preterm timepoint 1 = 23, preterm timepoint 2 = 97.

Figure S3 (relates to Figure 3). Bacterial species from shotgun metagenomic sequencing correlating with the first four orthogonal principal coordinates (PCo) calculated from the 16S-based data, showing the top 20 strongest correlations for each PCo. The % refers to the variance explained by each of the PCos. Red indicates positive and blue negative correlations between the PCo-s and species.

Figure S4 (relates to Figure 6). Species contribution to gut-brain modules.

(A) Correlation heatmap showing the Spearman rank correlation between the relative abundance of 25 most abundant species and gut-brain modules; modules are ordered by the mean relative abundance in the dataset. Red indicates positive and blue negative correlations.

(B) Relative abundance of the six gut-brain modules associated with MRI features stratified by species to show the contributions from known and unknown bacteria; for each module, only seven species with the highest abundance of that module are shown. Depth of the blue colour represents the mean relative abundance of the species in the dataset, with darker colours corresponding to higher overall relative abundance.

Figure S5 (relates to Figures 4-5). Representative brain maps.

Top panel: segmentation of the brain tissues of interest, overlaid on the Developing Human Connectome Project 40-week T2w template; middle panel: diffusion tensor imaging maps; bottom panel: neurite orientation dispersion and density imaging maps using the parallel diffusivity values for neonatal white matter (left) and grey matter (right). Maps are averaged over 20 random participants in this study.

### TABLES

Table S1 (relates to Table 1). Additional characteristics of the study group.

Table S2 (relates to Figure 2). MaAsLin2 results for the model testing for the effects of perinatal covariates in preterm samples at timepoint 1, adjusting for postnatal age at sample collection. ASVs present with at least 1% of abundance in at least 5% of samples were analysed.

Table S3 (relates to Figure 2). MaAsLin2 results for the model testing for the effects of perinatal covariates in preterm samples at timepoint 2, adjusting for degree of prematurity and GA at sample collection. ASVs present with at least 1% of abundance in at least 5% of samples were analysed.

Table S4 (relates to Figure 2). MaAsLin2 results for the model testing for the effects of perinatal covariates in preterm samples at timepoint 1, adjusting for postnatal age at sample collection. Species present with at least 1% of abundance in at least 5% of samples were analysed.

Table S5 (relates to Figure 2). MaAsLin2 results for the model testing for the effects of perinatal covariates in preterm samples at timepoint 2, adjusting for degree of prematurity and GA at sample collection. Species present with at least 1% of abundance in at least 5% of samples were analysed.

Table S6 (relates to Figure 2). Associations between perinatal variables and alpha diversity indices. P-values were adjusted for FDR using the Benjamini-Hochberg method. Sepsis, necrotising enterocolitis and bronchopulmonary dysplasia refer to diagnosis of these at any time during admission.

Table S7 (relates to Figures 4-6 and Table 1). Individual infant TP2 microbiome and brain MRI scan ages. There is a weak correlation between GA at TP2 sample and GA at MRI (Spearman rho = 0.122).

Table S8 (relates to Figure 4). Associations of global volumetric and microstructural MRI features with GA at birth and at scan. Results from linear regression model with each of the MRI feature as the outcome and GA at birth and at scan as the predictors. All values were scaled (z-transformed) before fitting the models, thus, the regression coefficients are in the units of standard deviations. cGM = cortical grey matter, dGM = deep grey matter, WM = white matter, CB = cerebellum, sulc = sulcal depth, GI = gyrification index, FA = fractional anisotropy, RD = radial diffusivity, NDI = neurite density index, ODI = orientation dispersion index, ISO = isotropic volume fraction, g = general factor.

Table S9 (relates to Figure 5). ASV-level analyses correlating brain microstructural features with the relative abundances of ASVs. Analyses were conducted using MaAsLin2, testing differences in ASVs present with at least 1% of abundance in at least 10% of samples (n=13 ASVs). Brain features were tested in separate models. Brain features were first adjusted for gestational age at birth and at MRI via linear regression, retaining the residuals. Then, MaAsLin2 models were performed for the microbiota composition table as the outcome and each residualised brain MRI feature as the predictor, adjusting for gestational age at faecal sample collection. FA = fractional anisotropy; NDI = neurite density index; ODI = orientation dispersion index; cGM = cortical grey matter; dGM = deep grey matter, g = general factor.

Table S10 (relates to Figure 5). Taxa-level analyses correlating brain microstructural features with the relative abundances of species. Analyses were conducted using MaAsLin2, testing differences in species present with at least 1% of abundance in at least 10% of samples (n=16 species). Brain features were tested in separate models. Brain features were first adjusted for gestational age at birth and at MRI via linear regression, retaining the residuals. Then, MaAsLin2 models were performed for the microbiota composition table as the outcome and each residualised brain MRI feature as the predictor, adjusting for gestational age at faecal sample collection. FA = fractional anisotropy; NDI = neurite density index; ODI = orientation dispersion index; cGM = cortical grey matter; dGM = deep grey matter, g = general factor.

Table S11 (relates to Figure 6). Module-level analyses correlating brain microstructural features with the relative abundances of gut-brain modules. Analyses were conducted using MaAsLin2, with default settings (testing differences in modules present in at least 10% of samples; n=34 modules). Brain features were tested in separate models. Brain features were first adjusted for gestational age at birth and at MRI via linear regression, retaining the residuals. Then, MaAsLin2 models were performed for the microbiota composition table as the outcome and each residualised brain MRI feature as the predictor, adjusting for gestational age at faecal sample collection. FA = fractional anisotropy; NDI = neurite density index; ODI = orientation dispersion index; cGM = cortical grey matter; dGM = deep grey matter, g = general factor.

