## Supplemental figures for "The neonatal gut microbiota: a role in the encephalopathy of prematurity"

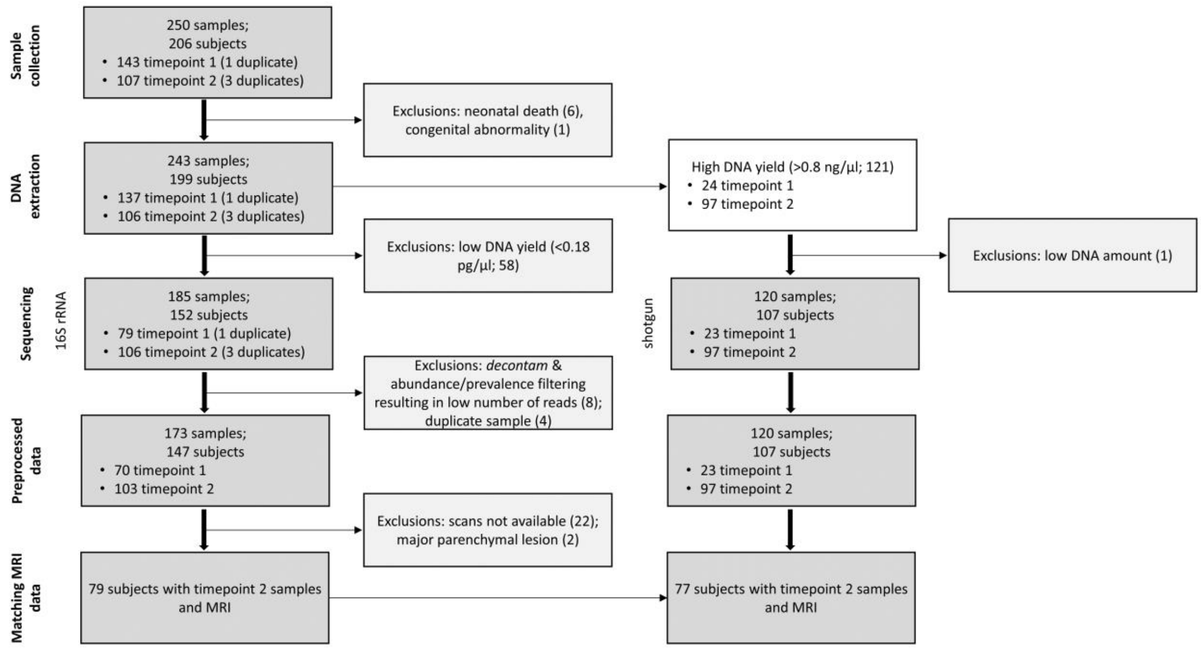


**Figure S1 (relates to Table 1).** Flowchart detailing the inclusion and exclusion of samples and participants in the study.

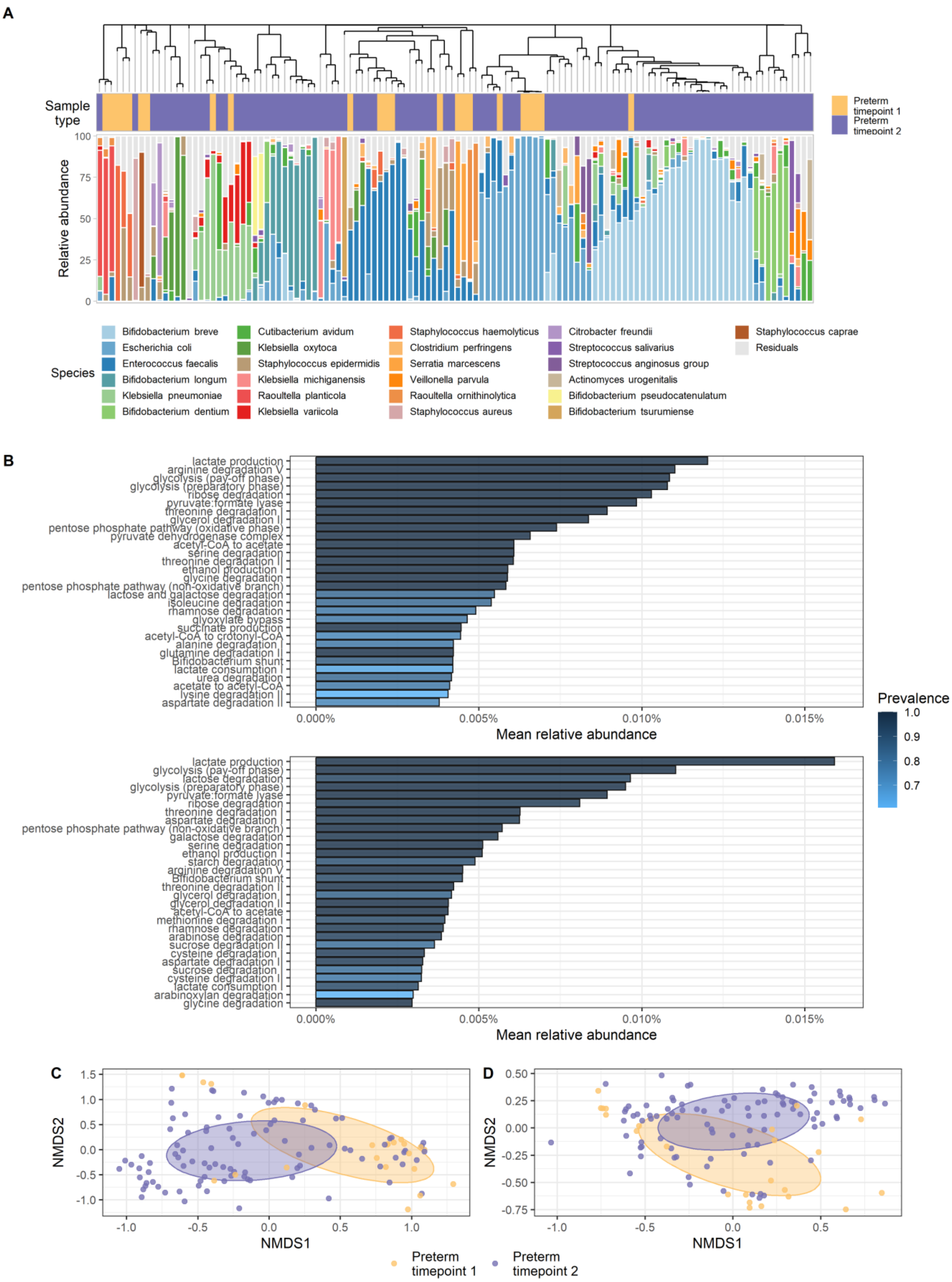


**Figure S2 (relates to Figure 1).** Overview of microbiome profiles in preterm neonates from shotgun metagenomic sequencing.

(A) Relative abundances of the 25 most abundant species identified across the dataset are visualised per sample. Samples are ordered based on hierarchical clustering of the Bray-Curtis dissimilarity matrix using complete linkage (see dendrogram).
(B) Mean relative abundances of the 30 most abundant gut metabolic modules in preterm infant stool at timepoint 1 (top) and 2 (bottom); bars are coloured by the prevalence of the modules at the two timepoints.
(C,D) Non-metric multidimensional scaling plot based on Bray-Curtis dissimilarity between samples at (C) species (PERMANOVA $R^2$ = 3.31%, p = 0.002) (D) and gut metabolic modules (PERMANOVA $R^2$ = 4.17%, p = $9.99 \times 10^{-4}$) level; data points and ellipses are coloured by sample type. The ellipses denote the standard deviation of data points belonging to each sample type, with the centre points of the ellipses calculated using the mean of the coordinates per group. 94/103 gut-brain modules were identified in the dataset.
Sample sizes: preterm timepoint 1 = 23, preterm timepoint 2 = 97.

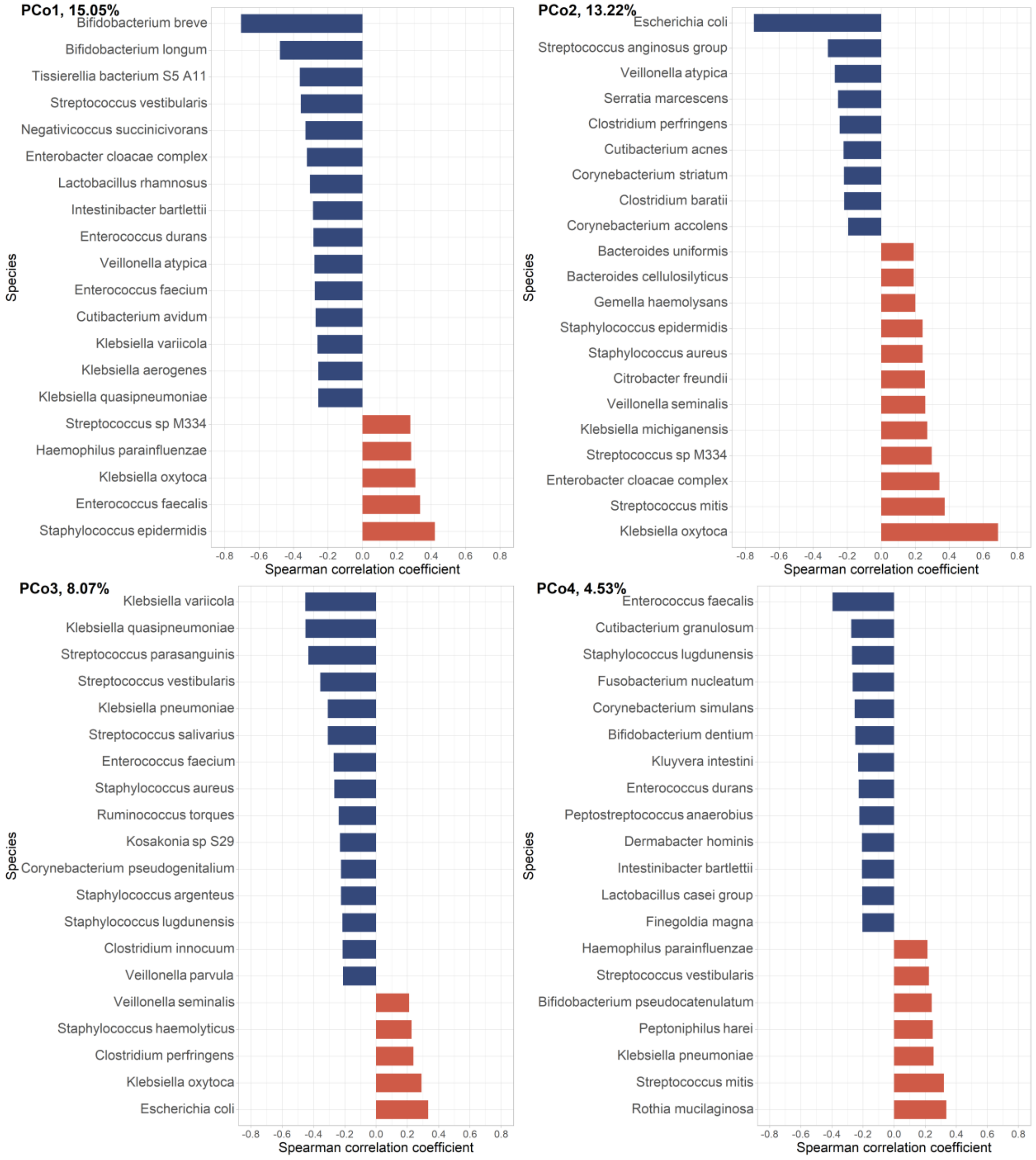


**Figure S3 (relates to Figure 3).** Bacterial species from shotgun metagenomic sequencing correlating with the first four orthogonal principal coordinates (PCo) calculated from the 16S-based data, showing the top 20 strongest correlations for each PCo. The % refers to the variance explained by each of the PCos. Red indicates positive and blue negative correlations between the PCo-s and species.

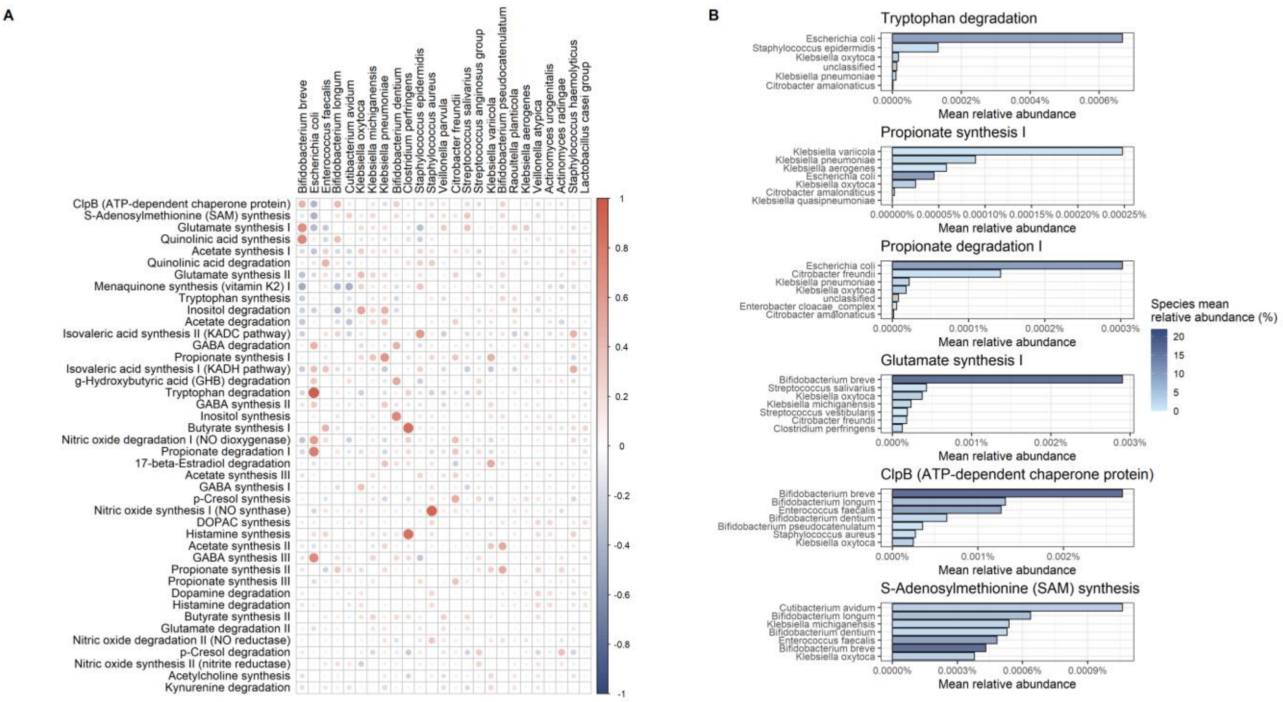


**Figure S4 (relates to Figure 6). Species contribution to gut-brain modules.**

(A) Correlation heatmap showing the Spearman rank correlation between the relative abundance of 25 most abundant species and gut-brain modules; modules are ordered by the mean relative abundance in the dataset. Red indicates positive and blue negative correlations.

(B) Relative abundance of the six gut-brain modules associated with MRI features stratified by species to show the contributions from known and unknown bacteria; for each module, only seven species with the highest abundance of genes present in that module are shown. Depth of the blue colour represents the mean relative abundance of the species in the dataset, with darker colours corresponding to higher overall relative abundance.

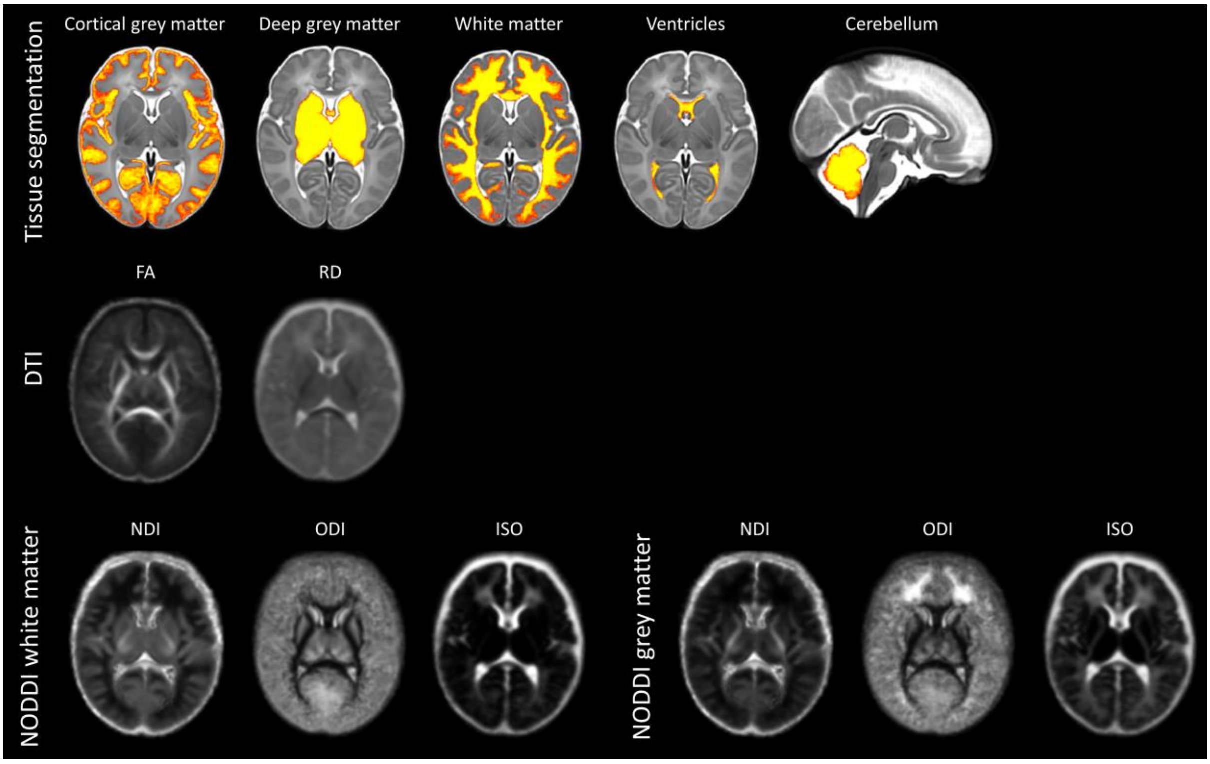


**Figure S5 (relates to Figures 4-5). Representative brain maps.**
Top panel: segmentation of the brain tissues of interest, overlaid on the Developing Human Connectome Project 40-week T2w template; middle panel: diffusion tensor imaging maps; bottom panel: neurite orientation dispersion and density imaging maps using the parallel diffusivity values for neonatal white matter (left) and grey matter (right). Maps are averaged over 20 random participants in this study.
